# The genomic landscape across 474 surgically accessible epileptogenic human brain lesions

**DOI:** 10.1101/2022.03.14.22271705

**Authors:** Javier A. López-Rivera, Costin Leu, Marie Macnee, Jean Khoury, Lucas Hoffmann, Roland Coras, Katja Kobow, Nisha Bhattarai, Eduardo Pérez-Palma, Hajo Hamer, Sebastian Brandner, Karl Rössler, Christian G. Bien, Thilo Kalbhenn, Tom Pieper, Till Hartlieb, Elizabeth Butler, Giulio Genovese, Kerstin Becker, Janine Altmüller, Lisa-Marie Niestroj, Lisa Ferguson, Robyn M. Busch, Peter Nürnberg, Imad Najm, Ingmar Blümcke, Dennis Lal

## Abstract

Understanding the exact molecular mechanisms involved in the etiology of epileptogenic pathologies with or without tumor activity is essential for improving treatment of drug-resistant focal epilepsy. Here, we characterize the landscape of somatic genetic variants in resected brain specimens from 474 individuals with drug-resistant focal epilepsy using deep whole-exome sequencing (>350×) and whole-genome genotyping. Across the exome, we observe a greater number of somatic single-nucleotide variants (SNV) in low-grade epilepsy-associated tumors (LEAT; 7.92±5.65 SNV) than in brain tissue from malformations of cortical development (MCD; 6.11±4 SNV) or hippocampal sclerosis (HS; 5.1±3.04 SNV). Tumor tissues also had the largest number of likely pathogenic variant carrying cells. LEAT had the highest proportion of samples with one or more somatic copy number variants (CNV; 24.7%), followed by MCD (5.4%) and HS (4.1%). Recurring somatic whole chromosome duplications affecting Chromosome 7 (16.8%), chromosome 5 (10.9%), and chromosome 20 (9.9%) were observed among LEAT. For germline variant-associated MCD genes such as *TSC2, DEPDC5*, and *PTEN*, germline SNV were frequently identified within large loss of heterozygosity regions, supporting the recently proposed ‘second hit’ disease mechanism in these genes. We detect somatic variants in twelve established lesional epilepsy genes and demonstrate exome-wide statistical support for three of these in the etiology of LEAT (e.g., *BRAF*) and MCD (e.g., *SLC35A2* and *MTOR*). We also identify novel significant associations for *PTPN11* with LEAT and NRAS Q61 mutated protein with a complex MCD characterized by polymicrogyria and nodular heterotopia. The variants identified in *NRAS* are known from cancer studies to lead to hyperactivation of NRAS, which can be targeted pharmacologically. We identify large recurrent 1q21-q44 duplication including *AKT3* in association with focal cortical dysplasia type 2a with hyaline astrocytic inclusions, another rare and possibly under-recognized brain lesion. The clinical genetic analyses showed that the numbers of somatic SNV across the exome and the fraction of affected cells were positively correlated with the age at seizure onset and surgery in individuals with LEAT. Finally, we report that identifying a likely pathogenic variant enabled us to refine or reclassify previous histopathological classifications *post hoc* in 20.5% of diagnoses. In summary, our comprehensive genetic screen sheds light on the genome-scale landscape of genetic variants in epileptic brain lesions, informs the design of gene panels for clinical diagnostic screening, and guides future directions for clinical implementation of epilepsy surgery genetics.

## Introduction

Drug-resistant epilepsies due to focal brain lesions represent a huge health burden and challenge for everyday clinical practice.^1^ Neurosurgical resection strategies have proven helpful in carefully selected patients, especially for brain lesions visible through magnetic resonance imaging (MRI) and confirmed by histopathology diagnosis.^2,3^ The most common types of epilepsy-associated brain lesions comprise hippocampal sclerosis (HS)^4^, low-grade epilepsy-associated developmental brain tumors (LEAT)^5,6^, such as ganglioglioma (GG) and dysembryoplastic neuroepithelial tumors (DNET), and malformations of cortical development (MCD), such as focal cortical dysplasia (FCD).^7^ Overall, these three most common categories accounted for more than 80% of the almost 10,000 patients who underwent epilepsy surgery and were collected in the European Epilepsy Brain Bank.^2^

Genetic factors have been associated with many common^8^ and rare epilepsies.^9–13^ Until recently, specific genes with variants of large effects have mainly been discovered in rare and severe forms of pediatric epilepsy that typically do not show structural abnormalities on MRI.^10–13^ In the last decade, more than 15 genes have been associated with somatic variants in epilepsy-associated MCD and LEAT.^5,14–16^ In contrast and to the best of our knowledge, the burden of somatic variants has not been systematically evaluated yet for HS. For MCD, somatic variants in genes encoding proteins of the canonical PI3K-AKT-MTOR pathway and germline loss-of-function variants—with or without co-occurring somatic losses of heterozygosity—in genes encoding proteins of the GATOR complex, a negative regulator of the PI3K-AKT-MTOR pathway, have been associated with FCD Type 2 and hemimegalencephaly (HME).^15–18^ In addition, the *SLC35A2* gene has been associated with the recently discovered mild MCD with oligodendroglial hyperplasia in epilepsy (MOGHE) disease entity.^19,20^ For LEAT, somatic variants that affect the RAS-RAF-MAPK pathway (i.e., *BRAF* and *FGFR1*) play a major role, with the *BRAF* variant V600E reported in 18–56% of GG and pathogenic variants in *FGFR1* in 58–82% of DNET.^5,21,22^ There is also emerging evidence for somatic variants in the RAS-RAF-MAPK pathway underlying MCD, with recent case studies reporting variants in *KRAS* associated with epilepsy-associated tumors and malformations.^23–25^

The search for actionable treatment targets in drug-resistant focal epilepsies pushed genetic studies of epilepsy-associated brain lesions into an emerging field. Most current studies utilize ultra-deep sequencing with coverages >1000x to identify low allelic fraction disease-causing single nucleotide variants (SNV) with high sensitivity and specificity. Due to the high coverage and correlated high sequencing cost, current studies typically focus on a small set of genes.^16,26^ Nevertheless, this approach has been very successful, leading to the discovery of 15 lesional epilepsy-associated genes.^5,14–16^ However, such candidate gene approaches did not interrogate the mutational signature for a structural lesion across the whole exome or genome, which is a typical study design in cancer research.^27,28^ As a result, the mutation rate and genetic architecture across epileptic brain lesion categories have not been systematically described. Subsequently, it is unclear whether MCD are single gene or oligogenic somatic disorders and whether somatic variation contributes to the development of HS. In addition, the role of genome-wide postzygotic copy-number variants (CNV) in different epileptic lesions has not been investigated. Recent evidence suggests that low-frequency mosaicism of variants in germline epilepsy-associated ion-channel encoding genes (e.g., *SCN8A*) can cause epilepsy in rodent models.^29^ However, it is unclear if somatic variants in these genes can cause focal epilepsy in humans. It is also unclear if variants in the 15 recently identified genes^5,14–16^ represent the most common genetic cause of lesional epilepsy as most previous studies were gene panel candidate studies or small-scale exome-wide studies, underpowered for statistical gene burden analyses across the exome.^16,26^

Here, we explored the exome-wide somatic variant burden and the genome-wide copy number variant burden of the three most common categories of epileptogenic brain lesions (i.e., LEAT, MCD, and HS). We analyzed differential variant burden across lesion categories, statistically identified novel epileptogenic pathology-associated genes, explored the role of driver mutations observed in cancer, and refined genotype-phenotype associations. The results of our study will shed light on the genetic architecture for a large number of epileptogenic brain lesions and inform the design of diagnostic genetic tests with reliably high sensitivity and specificity.

## Methods

### Study cohorts

Snap frozen surgical brain tissue samples obtained from 474 individuals with epilepsy-associated brain lesions (223 MCD, 154 LEAT, and 97 HS) were retrieved from the Cleveland Clinic Epilepsy Center biorepository (*n*=154) and European Epilepsy Brain Bank consortium (*n*=320). All studies were performed following institutional guidelines and regulations regarding research involving human subjects and approved by the ethics review boards of the Cleveland Clinic and the University of Erlangen, Germany. All patients underwent a comprehensive presurgical evaluation followed by a discussion at a multidisciplinary patient management conference where the surgical strategy was approved. Histopathological review of all resected brain tissue was performed and interpreted by board-certified clinical neuropathologists at the Cleveland Clinic in all patients and followed by a detailed re-review by experienced neuropathologists (I.B. and R.C.) using the ILAE classification scheme for FCD^7^, HS^4^, and the 4^th^ edition of the WHO classification of tumors of the central nervous system.^30^ Characteristics of our study cohorts are detailed in Table 1.

**Table 1:** Study cohort.

|  | **Total (n = 474)** | **CCF (n = 154)** | **EEBB (n = 320)** |
| --- | --- | --- | --- |
| **Hippocampal sclerosis (HS)** | **97** | **3** | **94** |
| Type 1 HS | 52 | 2 | 50 |
| Type 2 HS | 6 | 0 | 6 |
| Type 3 HS | 2 | 0 | 2 |
| HS (NOS) | 37 | 1 | 36 |
| **Malformation of cortical development (MCD)** | **223** | **111** | **112** |
| FCD 1 | 9 | 5 | 4 |
| FCD 1a | 8 | 4 | 4 |
| FCD 1b | 1 | 1 | 0 |
| MOGHE | 31 | 7 | 24 |
| FCD 2 | 111 | 46 | 65 |
| FCD 2a | 45 | 25 | 20 |
| FCD 2b | 65 | 21 | 44 |
| FCD 2 (NOS) | 1 | 0 | 1 |
| FCD (NOS) | 30 | 30 | 0 |
| HME | 7 | 1 | 6 |
| PMG | 10 | 3 | 7 |
| Other MCD | 25 | 19 | 6 |
| mMCD | 22 | 16 | 6 |
| NH | 3 | 3 | 0 |
| **Low-grade epilepsy-associated tumors (LEAT)** | **154** | **40** | **114** |
| Ganglioglioma | 83 | 8 | 75 |
| DNET | 24 | 5 | 19 |
| Other LEAT | 47 | 27 | 20 |
| Glioma | 16 | 0 | 16 |
| MVNT | 3 | 1 | 2 |
| LEAT (NOS) | 26 | 26 | 0 |
| Meningioangiomatosis | 2 | 0 | 2 |

CCF = Cleveland Clinic Foundation, EEBB = European Epilepsy Brain Bank, FCD = Focal cortical dysplasia, NOS = not otherwise specified, mMCD = mild MCD, MOGHE = mMCD with oligodendroglial hyperplasia in epilepsy, HME = Hemimegalencephaly, PMG = Polymicrogyria, NH = Nodular heterotopia, DNET = Dysembryoplastic neuroepithelial tumor, MVNT = Multinodular vacuolated neuronal tumor

Genomic DNA was extracted from fresh-frozen brain tissue for all 474 individuals. The DNeasy Blood and Tissue Kit (Qiagen) was used for DNA extraction from fresh frozen brain samples according to the manufacturer’s protocol.

### Somatic and germline SNV calling

Deep whole-exome sequencing (>350x WES) of all samples in this study was performed using Agilent SureSelect Human All Exon V7 enrichment and paired-end reads (151bp) Illumina sequencing. All paired-end FASTQ files were aligned to the GRCh37/hg19 human reference genome, including the hs37d5 decoy sequence using BWA-MEM^31^, following GATK best practices.^32^ We used MuTect2^33^ to generate a panel of normals (PoN) from 21 additional surgically resected and sequenced brain samples to exclude sequencing artifacts and germline variants. All 21 samples included in the PoN underwent surgery for epilepsy and were histopathologically confirmed as non-lesional or due to an external insult (glial scar or encephalitis). We then called somatic SNV, insertions, and deletion polymorphisms (indels) using MuTect2^33^ in conjunction with the PoN. We only retained somatic variants with a maximum minor allele frequency (MAF) <10^−5^ in the Genome Aggregation Database (gnomAD). Variants within segmental duplication regions or nondiploid regions were removed. Low-quality calls flagged by MuTect2 with ‘t_lod_fstar’, ‘str_contraction’, and ‘triallelic_site’ were removed. Finally, we excluded somatic indels within RepeatMasker^34^ or simple repeat regions.

We then used MosaicForecast^35^ to perform read-based phasing and identify high-confidence somatic mosaic variants from all called variants. MosaicForecast is a machine-learning-based method optimized to detect somatic mutations without a matched reference tissue. Briefly, the method trains a random forest model on read-based features from phased variants and has been shown to reliably identify somatic mosaic mutations with variant allelic fractions (VAF) as low as 2%. Therefore, the method has the sensitivity to detect variants present in as few as 4% of cells in a given sample of brain tissue.^35,36^ After all training and filtration, we identified 217,560 putative somatic mosaic SNV across all samples. We then filtered all exonic or splice site variants with a VAF between 0.02 and 0.40 and an alternative read depth ≥3. Only nonrecurring mutations within the cohort were considered, except for variants in 15 genes that have been previously associated with epileptogenic lesions: *MTOR, SLC35A2, AKT3, PIK3CA, RHEB, TSC1, TSC2, NPRL2, NPRL3, DEPDC5, PTEN, BRAF, FGFR1, MYB*, and *MYBL1*.^5,14–16^ Lastly, to ensure that no potentially meaningful variants in established lesional epilepsy genes were mistakenly filtered out, we used the Integrated Genomics Viewer (IGV)^37^ browser to visually examine all somatic variants in established epileptic lesion genes called by Mutect2-PoN, but judged as potential artifacts by MosaicForecast. However, such variants were not included in the somatic variant enrichment analyses and were only included when reporting the final yield from our screen.

Finally, we used GATK’s HaplotypeCaller (GATK 4.1.9.0)^38^, following GATK best practices^39^, to identify germline variants in lesional epilepsy-associated genes in negative regulators shown to act through a germline or two-hit mechanism (i.e., *TSC1, TSC2, DEPDC5, NPRL2, NPRL3*, and *PTEN*).^5,14–16^ We also used GATK’s HaplotypeCaller to identify somatic variants of high allelic fraction (VAF>0.30) in other lesional epilepsy-associated genes (i.e., *MTOR, SLC35A2, AKT3, PIK3CA, RHEB, BRAF, FGFR1, MYB*, and *MYBL1*)^5,14–16^ that MuTect2 with PoN or MosaicForecast may have missed. The resulting variants were filtered for variants with **(1)** phred quality score (QUAL)<30, **(2)** genotype quality (GQ)<99, **(3)** sample read depth (DP)≥30, **(4)** max DP<1,000, and **(5)** GATK truth sensitivity tranche >99.5% for single nucleotide variants (SNV) and >95% for indels and were not included in the somatic variant enrichment analyses (Method: “Exome-wide statistical identification of genes associated with epileptogenic brain lesions”).

### Somatic CNV and CNN-LOH calling

A total of 688,032 single nucleotide polymorphisms (SNPs) were genotyped for all samples of this study using the Global Screening Array with Multi-disease drop-in (GSA-MD v1.0; Illumina, San Diego, CA, USA). The SNP data was used to detect somatic CNV using MoChA. MoChA is available as a bcftools^40^ extension that uses phased VCF files with B Allele Frequency (BAF) and Log R Ratios (LRR) to identify somatic CNV and copy number neutral loss of heterozygosity (CNN-LOH). MoChA employs a three-state hidden Markov model (HMM) to capture somatic-CNV-induced deviations in allelic balance (|ΔBAF|) at heterozygous sites. After affine-normalization and GC wave-correction, the BAF and LRR values were transformed as described elsewhere.^41^ Autosomes and sex chromosomes were separately considered. Before phasing with Eagle v2^42^, we generated a list of variants that was excluded from modeling by both Eagle and MoChA, using the following parameters: **(1)** segmental duplications with low divergence (<2%), **(2)** high levels of missingness (>2%), **(3)** variants with excess heterozygosity (*P*<10^−6^), and **(4)** variants that unexpectedly correlate with sex (*P*<10^−6^). After somatic CNV calling, we removed samples based on the following quality control (QC) parameters: **(1)** call rates <0.97, and **(2)** baf_auto>0.3. We then removed variants based on the following QC parameters: **(1)** lod_baf_phase<10 unless the somatic CNV was larger than 5Mbp (or 10Mbp if they span the centromere), **(2)** CNV calls flagged as germline CNV, and **(3)** possible constitutional duplications with length>10Mb and either LRR>0.35 or LRR>0.2 and |ΔBAF|>0.16 or length<10Mb and either LRR>0.2 or LRR>0.1 and |ΔBAF|>0.1.^43^ The estimation of the allelic fraction for each somatic CNV or CNN-LOH by MoChA has been detailed elsewhere.^44^ All somatic CNV and CNN-LOH were examined visually by plotting the LRR, BAF, and phased BAF values with MoChA.

### Variant annotation and assessment of deleteriousness

We applied different strategies for the identified CNV and SNV to assess the likelihood of a deleterious effect on disease-relevant loci or genes. We used ANNOVAR^45^ with custom databases to perform variant annotation. The deleteriousness of SNV used to characterize the genetic architecture of the different brain lesion categories was assessed based on two filters: **(1)** variant type and population frequency and **(2)** predicted variant deleteriousness. The frequency filter was based on the variant not being present at an AF>10^−5^ in the gnomAD^46^ database. From the remaining variants, we selected only variants with a high confidence prediction to be deleterious using the following criteria: **(1)** loss-of-function (LoF) variants ranked in the top 1% most deleterious variants in the human genome according to the Combined Annotation Dependent Depletion score (CADD, scaled CADD phred score ≥ 20)^47^ found in established lesional epilepsy genes^5,14–16^ or highly loss-of-function (LoF) intolerant genes (genes with a gnomAD LoF Observed/Expected Upper bound Fraction LOEUF≤0.35)^46^ and **(2)** missense variants ranked in the top 1% most deleterious variants in the human genome genome (scaled CADD phred score ≥20)^47^ in established lesional epilepsy genes or found in missense-constrained sites (MPC score ≥2)^48,49^ of missense intolerant genes (gnomAD missense Z-score >3.09, corresponding to *P*<10^−3^).^46^ The supporting aligned reads of all reported variants were visually inspected using the IGV browser.^37^

For somatic CNV, we considered all somatic CNV or CNN-LOH that had a size >1Mbp and affected known cancer driver genes or genes associated with lesional epilepsies (i.e., *MTOR, SLC35A2, AKT3, PIK3CA, RHEB, TSC1, TSC2, NPRL2, NPRL3, DEPDC5, PTEN, BRAF, FGFR1, MYB*, and *MYBL1*)^5,14–16^ to be deleterious.

### Exome-wide statistical identification of genes associated with epileptogenic brain lesions

To detect genes under evolutionary mutational selection, we employed a Poisson-based dN/dS model using dNdScv.^50^ This model tests the normalized ratio of nonsynonymous (missense, nonsense, and splicing) over background (synonymous) mutations while correcting for sequence composition and mutational signatures. This method has been shown to reliably identify genes under positive selection in cancer and normal tissues.^50^ A global false discovery rate adjusted *P*-value *q*≤0.1 was used to identify statistically significant nonsynonymous variant-enriched genes and an unadjusted *P*≤0.005 to identify nominally significant nonsynonymous variant-enriched genes. Genes that showed a suggestive enrichment of nonsynonymous variants were only considered when their biological role and known disease associations matched the associated phenotype. To improve the quality of our results, the reads for all variants in genes identified by dNdScv were visually inspected using the IGV browser^37^ to assess variant quality. Low-quality variants were flagged as potential artifacts, and genes with only low-quality variants were excluded.

### Statistical analysis

All statistical analyses and filtering were done using R. We used the Chi-square, Fisher’s exact, or Wilcoxon rank-sum test where appropriate. Correlation statistics were generated with Pearson’s correlation.

### Data availability

All somatic variant calls are available from the supplementary material. All code is available at https://github.com/LalResearchGroup. Additional data are available from the corresponding author on reasonable request.

## Results

### Cohort overview

We extracted DNA from snap-frozen brain tissue samples of 474 individuals with drug-resistant epilepsy who underwent epilepsy surgery and were diagnosed with histopathologically confirmed HS, MCD, or LEAT (Table 1). HS and LEAT samples primarily originated from the temporal lobe, whereas MCD samples were primarily derived from extratemporal resections and had the largest proportion of multilobular involvement (Table 2). The average age at seizure onset for MCD patients was significantly earlier than that of HS and LEAT (MCD = 6.3 years, HS = 13.73 years, LEAT = 13.14 years; Wilcoxon rank-sum test: MCD vs. HS *P*=1.17×10^−7^, MCD vs. LEAT *P*=2.27×10^−10^). Conversely, the average age at surgery for HS patients was significantly later than that of MCD and LEAT (HS = 39.13 years, MCD = 18.48 years, LEAT = 21.82 years; Wilcoxon rank-sum test: HS vs. MCD *P*=1.02×10^−21^, HS vs. LEAT *P*=9.59×10^−15^, MCD vs. LEAT *P*=0.17). Lastly, individuals with HS had a longer duration of epilepsy before undergoing surgery, followed by individuals with MCD, and with LEAT (HS = 25.81, MCD = 12.24, LEAT = 8.53 years; Wilcoxon rank-sum test: HS vs. MCD *P*=4.33×10^−13^, HS vs. LEAT *P*=6.63×10^−19^, MCD vs. LEAT *P*=1.31×10^−3^).

**Table 2:**
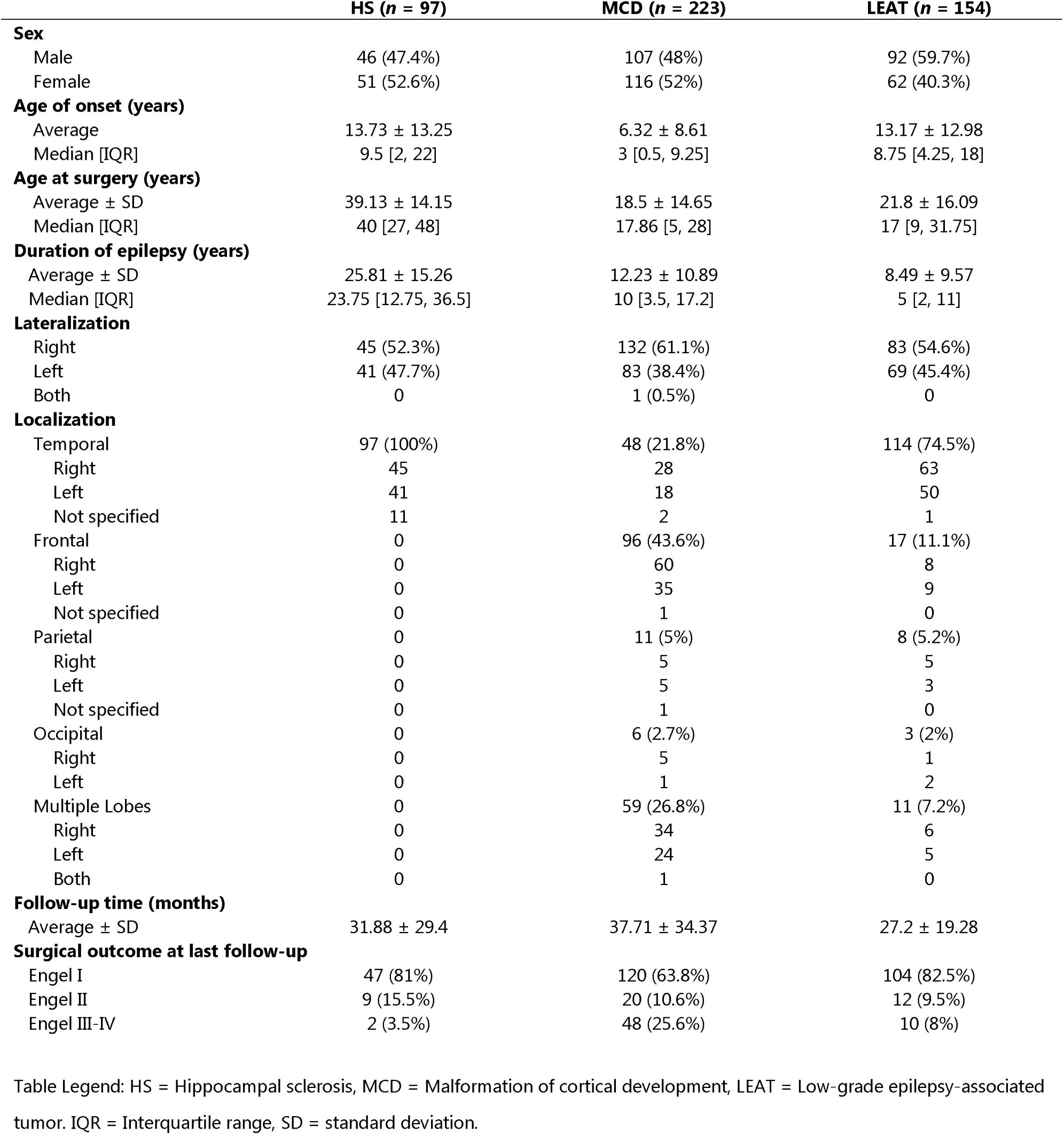
Clinical characteristics of the study cohort.

### Somatic SNV profiles differ across the major categories of epilepsy-associated brain lesions

Deep whole-exome sequencing achieved an average mean target coverage of 364x across all 474 samples. DNA quality and experimental performance across pathology groups were similar, with no differences in mean coverage among the three phenotype groups (Figure 1A). On average, the number of somatic SNV carried per sample was higher in LEAT (7.92±5.65) and MCD (6.11±4) than in HS (5.1±3.04, Figure 1B). From a total of 3,078 somatic SNV, 172 SNV affecting 141 individuals were bioinformatically classified as potentially deleterious. These included 128 missense variants and 44 protein-truncating variants. Overall, a higher proportion of LEAT (48.1%, 74/154) had at least one potentially deleterious SNV, compared to MCD (26.9%, 60/223) and HS (6.2%, 6/97, Figure 1C). The average variant allelic fraction (VAF) of somatic SNV was 1.18-fold higher in LEAT (average VAF=0.078) than in MCD (average VAF=0.066) and 1.26-fold higher than in HS (average VAF=0.062). MCD and HS had similar VAF distributions (Figure 1D). When considering only potentially deleterious variants, the average VAF in LEAT (average deleterious VAF=0.108) was 1.63-fold higher than in MCD (average deleterious VAF=0.066) and 1.22-fold higher than in HS (average deleterious VAF=0.088, Figure 1D). Overall, MCD and HS showed similar somatic SNV profiles, although MCD carried a nominally significant higher number of somatic SNV per sample and a larger proportion of samples with at least one deleterious missense SNV than HS (Figures 1B and 1C).

**Figure 1:**
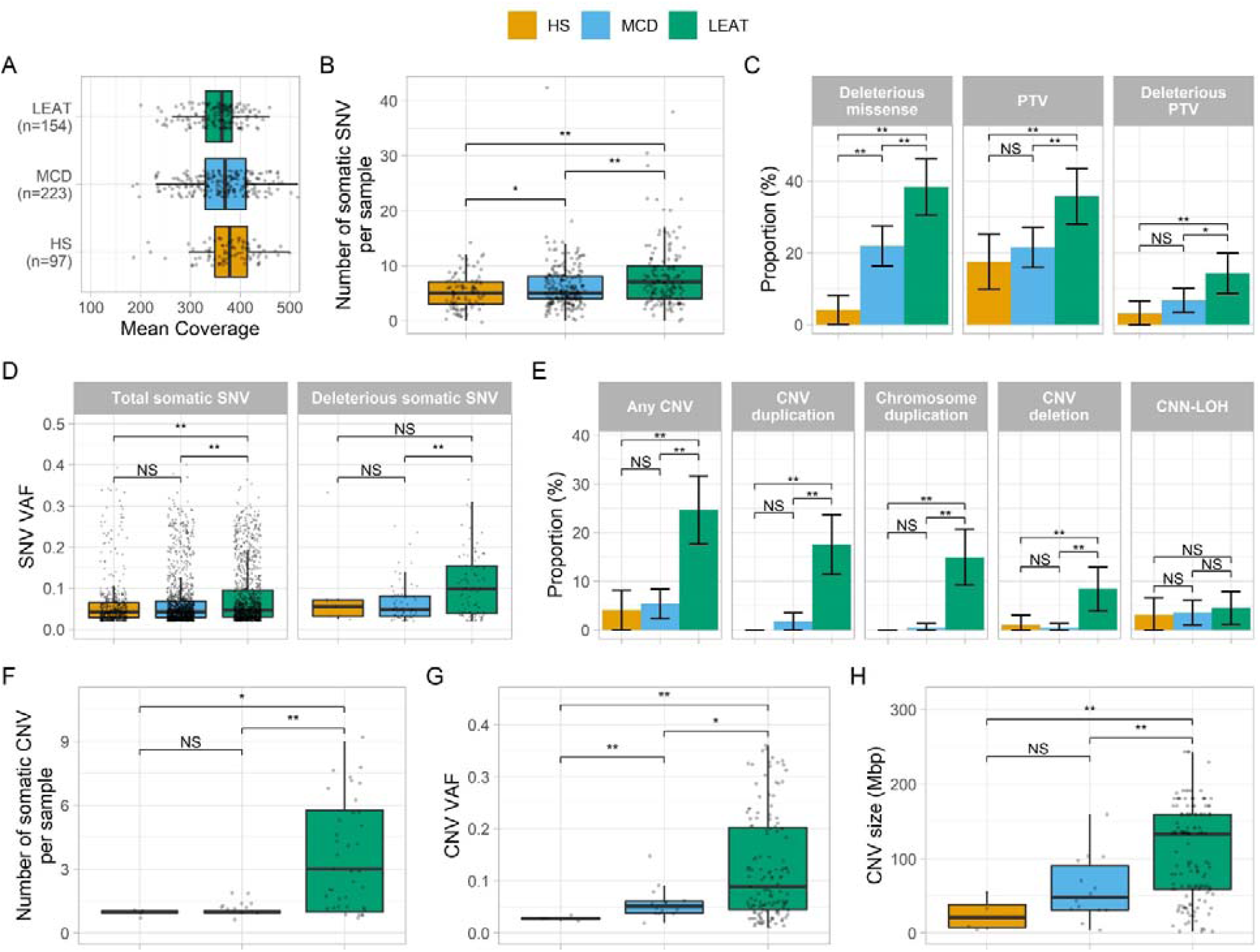
Different somatic mutation profiles observed across major categories of epilepsy-associated brain lesions. **(A)** Median coverage is similar across major lesion categories. (**B)** LEAT and MCD have a higher number of somatic SNV per sample than HS. (**C)** LEAT have a higher proportion of samples with at least one deleterious variant than HS and MCD. (**D)** The average variant allelic fraction (VAF) for total somatic SNV is higher in LEAT. Potentially deleterious variants (missense variants with MPC>2 and truncating variants in LoF-intolerant genes with LOEUF<0.35) in LEAT have a higher VAF than those in HS and MCD. (**E)** LEAT have a higher proportion of samples with at least one CNV than MCD and HS. (**F)** LEAT samples with somatic CNV have significantly more somatic CNV per sample than MCD/HS. (**G)** LEAT CNV occur at significantly higher VAF than MCD/HS. (**H)** LEAT have larger CNV than MCD/HS. HS = Hippocampal sclerosis, MCD = Malformations of cortical development, LEAT = Low-grade epilepsy-associated tumors, SNV = Single nucleotide variant, PTV = Protein-truncating variant, CNV = Copy-number variant, CNN-LOH = Copy number neutral loss of heterozygosity, VAF = Variant allelic fraction, * = nominally significant difference (*P*<0.05 before multiple-testing correction), ** = significant difference (*P*<0.05 after multiple-testing correction), NS = Not significant.

### Somatic CNV profiles differ across the major categories of epilepsy-associated brain lesions

After quality control, we identified a total of 153 large CNV and CNN-LOH across 56 samples: 105 duplications (85 whole chromosome gains), 27 deletions (5 whole chromosome losses), and 21 copy-number-neutral losses of heterozygosity (CNN-LOH). LEAT had the highest proportion of samples with one or more somatic CNV or CNN-LOH (38/154, 24.7%), followed by MCD (12/223, 5.4%) and HS (4/97, 4.1%; Figure 1E). LEAT were also enriched for CNV duplications and deletions compared to MCD and HS, whereas CNN-LOH were not distributed differently across all three categories. Among LEAT, somatic duplications were the most frequently observed type of CNV, with 71% (27/38) of CNV or CNN-LOH-positive samples carrying one or more somatic duplications. Somatic whole chromosome duplications were the major type of duplications among LEAT (82/101 LEAT gain CNV, 81.2%, Figure 1E). Conversely, somatic CNN-LOH were predominant among MCD and HS (8/12 CNV-positive MCD, 66.7%; 3/4 CNV-positive HS, 75%).

Most somatic CNV-positive LEAT had multiple somatic CNV (27/38, 71%), whereas only two (2/12, 16.7%) CNV-positive MCD had multiple somatic CNV or CNN-LOH (Figure 1F). No CNV-positive HS sample carried multiple somatic CNV or CNN-LOH. The highest average VAF of somatic CNV and CNN-LOH was identified in LEAT (average VAF=0.125), followed by MCD (average VAF=0.058), and HS (average VAF=0.028, Figure 1G). Lastly, LEAT samples carried larger somatic CNV and CNN-LOH (average size = 108.6 Mbp) than MCD (average size = 59.6 Mbp) and HS (average size = 25.3 Mbp, Figure 1H). Overall, LEAT displayed the most distinct CNV profile; they had a larger proportion of samples with CNV or CNN-LOH, were enriched for duplications and deletions, tended to carry more CNV per sample, and had larger CNV affecting a higher number of cells.

### The number of somatic SNV and the fraction of mutated tumor cells are associated with age at seizure onset and age at surgery in LEAT

For LEAT, we identified positive correlations between the number of somatic SNV per sample and later age at seizure onset and older age at surgery. (Pearson’s correlation; LEAT total somatic SNV & age at surgery: *r*=0.26, *P*=1.16×10^−3^; LEAT total somatic SNV & age at onset: *r*=0.22, *P*=9×10^−3^). Similarly, we also identified positive correlations between higher somatic VAF (a proxy marker for the number of mutated cells) and later age at seizure onset and older age at surgery for LEAT (Pearson’s correlation; LEAT somatic VAF & age at onset: *r*=0.15, *P*=1.61×10^−7^; LEAT somatic VAF & age at surgery: *r*=0.11, *P*=3.55×10^−5^). This signal was driven primarily by the somatic SNV VAF, with the VAF of the CNV and CNN-LOH having little effect (Pearson’s correlation; LEAT somatic SNV VAF & age at surgery: *r*=0.16, *P*=1.47×10^−8^; LEAT somatic SNV VAF & age at onset: *r*=0.18, *P*=1.44×10^−9^; LEAT somatic CNV VAF & age at surgery: *r*=-0.2, *P*=0.0176; LEAT somatic CNV VAF & age at onset: *r*=0.01, *P*=0.95). We did not identify any similar correlations between genetic architecture and clinical variables for MCD and HS.

### Unbiased somatic variant burden analysis confirms previously-reported and discovers novel lesional epilepsy-associated genes

We performed the first exome-wide burden analysis for a lesional epilepsy cohort (HS, MCD, and LEAT) to identify genes under positive mutational selection with significant enrichment of somatic SNV in any of the three main lesion categories. In line with previous results from candidate gene studies, we identified four genes with significant somatic SNV burden for LEAT and MCD and no genetic associations with HS (Supplementary Table 1, Figure 2A). For LEAT, the gene with the most significant exome-wide somatic SNV burden was *BRAF*, which represents the most well-established previously reported LEAT gene and is primarily associated with ganglioglioma.^5,51^ The second gene identified with a significant variant enrichment was *PTPN11*, which has not been previously associated with LEAT. The *PTPN11* gene encodes Protein Tyrosine Phosphatase Non-Receptor Type 11, an upstream regulator of the RAS/MAPK and mTOR signaling pathways. Activating mutations in *PTPN11* have been shown to cause Noonan Syndrome (a congenital developmental syndrome), play a role in tumorigenesis, and affect the development of white matter microstructure in humans.^52–54^ For MCD, our burden analysis identified *SLC35A2* and *MTOR*, which have been previously reported in multiple candidate gene studies or descriptive exome analysis screens as the two most frequently mutated genes in mild malformation of cortical development with oligodendroglial hyperplasia in epilepsy (MOGHE) and FCD type 2, respectively.^14^ However, we are the first to use an unbiased statistical approach to show that variants in these genes are a major genetic cause of MCD entities.

**Figure 2:**
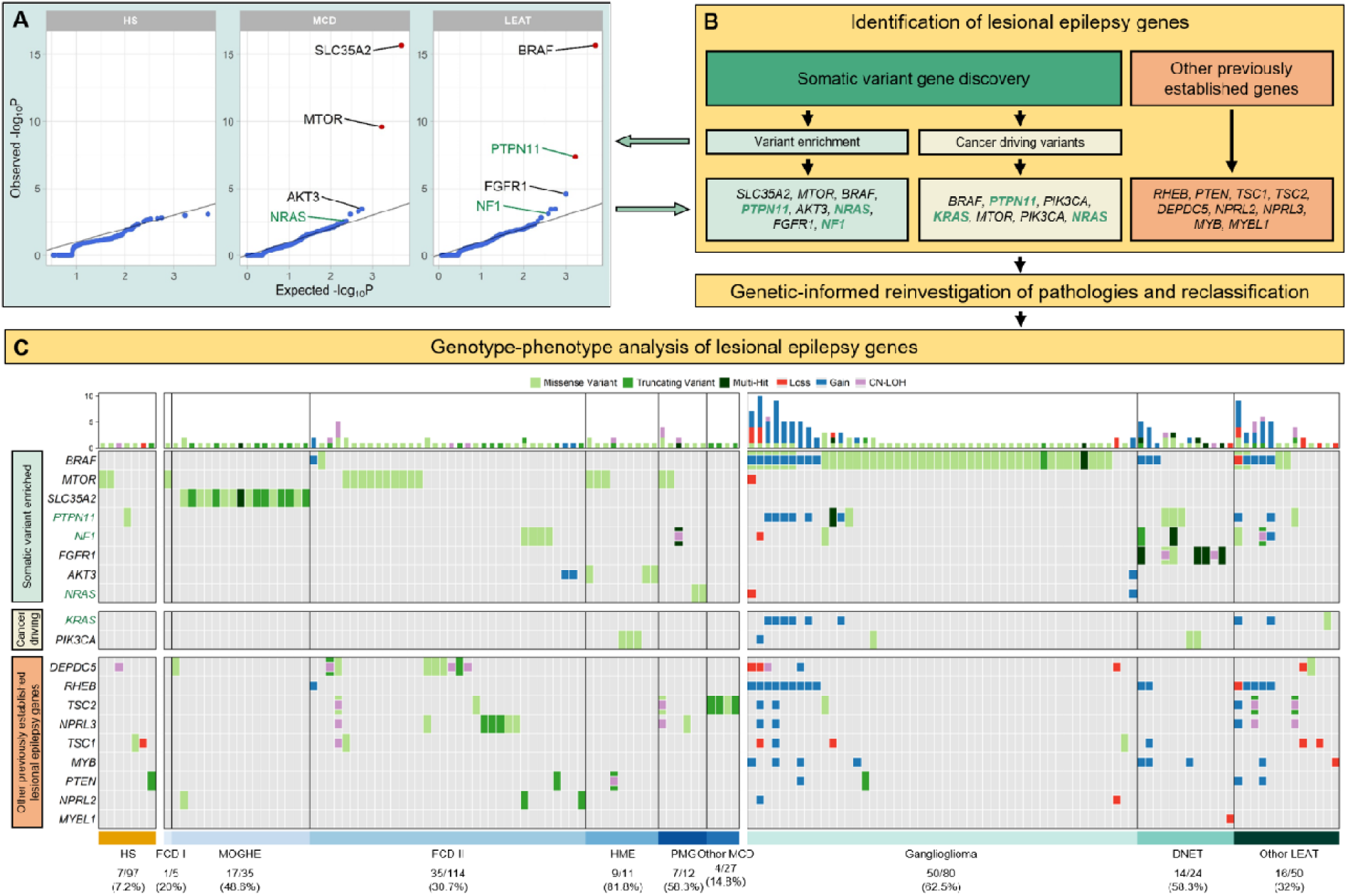
Characterization and histopathological annotation of genes associated with epileptogenic lesions. **(A)** Quantile-Quantile plots of the gene burden analysis results. Highlighted genes represent identified genes of interest (see methods) with novel genetic associations and candidate genes highlighted in green text. Red points indicate genes with statistically significant somatic variant enrichment (false discovery rate *q*<0.1). (**B)** Flow diagram for identifying genes of interest associated with lesional epilepsies and genotype-phenotype relationships. Novel genetic associations and candidate genes are highlighted in green text. (**C)** Somatic mutation profiles for 151 mutation-positive epileptogenic lesions. Each column represents an individual sample with colored rectangles highlighting identified variants across 19 genes associated with lesional epilepsies. Larger rectangles represent SNV and smaller squares represent CNV or CNN-LOH. Novel genetic associations and candidate genes are highlighted in green. CNN-LOH = Copy number neutral loss of heterozygosity, HS = Hippocampal sclerosis, MCD = Malformation of cortical development, LEAT = Low-grade epilepsy-associated tumor, FCD = Focal cortical dysplasia, mMCD = mild MCD, MOGHE = mMCD with oligodendroglial hyperplasia in epilepsy, HME = Hemimegalencephaly, PMG = Polymicrogyria, GG = Ganglioglioma, DNET = Dysembryoplastic neuroepithelial tumor.

### Identification of additional lesional epilepsy genes with suggestive statistical support and high biological plausibility

The previous statistical analysis could identify and validate the most frequently mutated lesional epilepsy genes that remained significant after controlling for the false discovery rate (*q*≤0.1). Next, we investigated variants in genes with suggestive variant enrichment (*P*<5×10^−3^) for association with LEAT or MCD by considering additional evidence criteria (see Methods and Supplementary Table 1). We also screened our cohort for variants in 579 high-confidence cancer driver sites, previously identified in 53 genes (Supplementary Table 2).^55^ The genes *FGFR1, NF1, PIK3CA*, and *KRAS* had additional evidence for a role in the LEAT etiology (Figure 2A). *FGFR1* and *NF1* had nominally significant variant enrichment and strong evidence in the literature based on patient reports with similar phenotypes (Supplementary Table 1).^5,21,22^ We also identified functionally validated cancer-driving variants in *PIK3CA* and *KRAS*, both of which are established tumor or growth disorder genes^56,57^ (Supplementary Table 2). The genes *AKT3, NRAS*, and *PIK3CA* had nominally significant mutational burden and/or variants in cancer-driving sites and met additional disease-association criteria for a role in the etiology of MCD (Supplementary Tables 1 and 2). *AKT3* and *PIK3CA* are well-established MCD genes and the identified variants have been molecularly characterized as activating variants.^58–60^ Additionally, we identified a novel association of the *NRAS* gene with MCD (Figure 3A). *NRAS* is a key regulator of cell proliferation, differentiation, and growth as an upstream regulator of the RAS-RAF-MAPK and PI3K-AKT-MTOR pathways.^61^ The variants driving the enrichment occurred at the previously established *NRAS* Q61 cancer-driving site (Supplementary Table 2). ^55,62^

**Figure 3:**
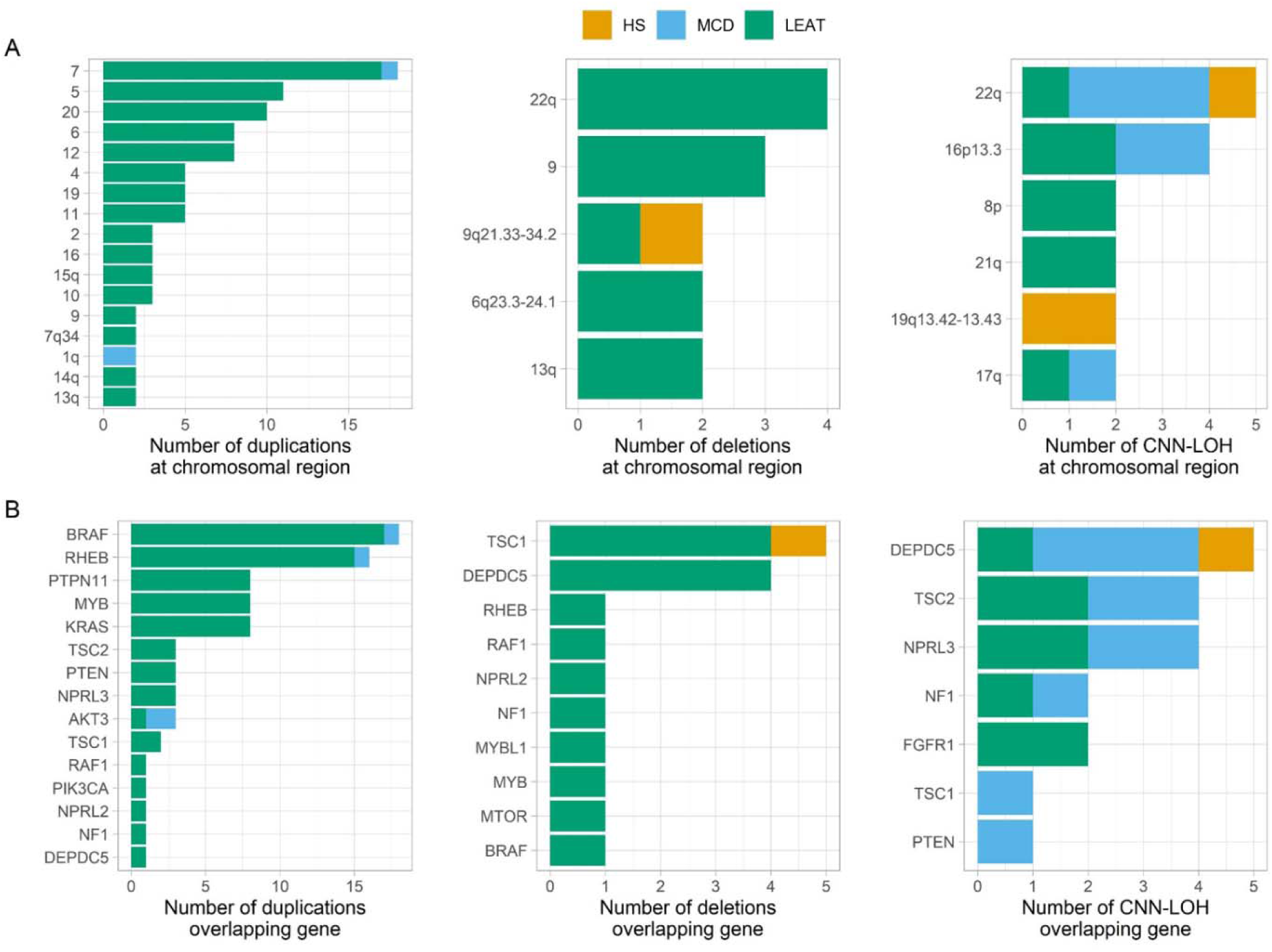
Recurring somatic CNV and CNN-LOH regions often overlap with lesional epilepsy genes. **(A)** Recurring regions of overlap among somatic copy number variants (CNV) and copy number neutral losses of heterozygosity (CNN-LOH). Smaller regions are identified based on included cytobands, CNV or CNN-LOH affecting >80% of a chromosomal arm are identified by the chromosomal arm, and CNV affecting the entire chromosome are identified by the chromosome number. (**B)** Detected CNV and CNN-LOH often overlap established and newly identified lesional epilepsy genes. HS = hippocampal sclerosis, MCD = malformation of cortical development, LEAT = Low-grade epilepsy-associated tumor

### Large somatic chromosomal alterations in lesional epilepsies frequently include established lesional epilepsy-associated genes

We identified several somatic CNV patterns and recurring regions across the lesion categories represented in our cohort. In LEAT specifically, the most frequently recurring duplicated regions were whole chromosome duplications of chromosome 7 (17/101, 16.8%), chromosome 5 (11/101, 10.9%), and chromosome 20 (10/101, 9.9%; Figure 3A). We also identified a recurring CNV gain of the q-arm of chromosome 1 in our MCD cohort (n=2). Duplications of chromosome 1q have been previously identified in cases of macrocephaly, megalencephaly, polymicrogyria, and focal cortical dysplasia.^63–65^ The most frequently recurring regions of CNN-LOH mapped to the longer q-arm of chromosome 22 (5/21, 23.8%) and chromosome 16p13.3 (4/21, 19%; Figure 3A). These regions were most prevalent in MCD, but were also identified in HS or LEAT. A single recurring region of CNN-LOH was unique to HS, which mapped to the q-arm of chromosome 19 (19q13.42-13.43).

To pinpoint the putative causative genes, we investigated whether the identified CNV and CNN-LOH overlapped with previously established lesional epilepsy genes. We observed that 68.4% of LEAT, 83.3% of MCD, and 50% of HS with a somatic CNV or CNN-LOH carried at least one chromosomal alteration affecting one of the 15 established lesional epilepsy genes^5,14–16^ (Figure 3B). The most frequently recurring CNV gain (chromosome 7) overlapped with the most frequently mutated gene in lesional epilepsies (*BRAF*). Additionally, the two most frequently recurring regions of CNN-LOH included *DEPDC5* (22q) or *TSC2* (16p13.3), both established lesional epilepsy genes that require a somatic second hit such as a CNN-LOH to cause disease.

### Genetic findings-driven histopathology reclassification and genotype-phenotype analyses

In total, we identified 19 genes of interest either through our somatic variant gene discovery analysis (*SLC35A2, MTOR, BRAF, PTPN11, AKT3, NRAS, FGFR1, NF1, PIK3CA*, and *KRAS*) or previously reported as genes associated with epileptogenic brain lesions (*RHEB, PTEN, TSC1, TSC2, DEPDC5, NPRL2, NPRL3, MYB*, and *MYBL1*).^5,14–16^ Across our entire cohort, we identified 151 samples that carried at least one SNV, CNV, or CNN-LOH affecting one of these genes (31.4% of cohort; 7.2% of HS, 31.8% of MCD, and 51.9% of LEAT; Figure 2C; Supplementary Tables 4 and 5). In addition to the evaluation upon enrollment, we performed an in-depth *post hoc* histopathological review of all mutation-positive samples in the context of their genetic diagnosis. From the 151 reviewed cases, we identified 31 (20.5%) instances in which the inclusion of a genetic diagnosis alongside an in-depth review led to either reclassification or refinement of the initial histopathological diagnosis (Supplementary Table 3). A conclusive diagnosis was determined for all 12 mutation-positive patient samples previously diagnosed as FCD or LEAT not otherwise specified (NOS) upon genetics-informed review. These challenging cases where a conclusive histopathological diagnosis could not be specified composed over a third of reclassified samples (12/32, 37.5%). In addition, the genetic information guided histopathological reclassification for 40.8% (29/71) of MCD and 2.7% (2/74) of LEAT samples (Supplementary Table 3).

Following histopathological review, the genes with the most variants (SNV, CNV, or CNN-LOH) and primarily homogeneous histopathology included: *BRAF* (52/67 in ganglioglioma), *SLC35A2* (17/17 in MOGHE), *FGFR1* (14/14 in DNET), and *MTOR* (13/19 in FCD 2b or HME). By considering known pathogenic pathways, we identified 74 samples (64 LEAT, 9 MCD, and one HS) with variants affecting genes in the RAS-RAF-MAPK pathway (*FGFR1, PTPN11, KRAS, NRAS, NF1*), 64 samples (32 MCD, 27 LEAT, and five HS) with variants in genes of the PI3K-AKT-MTOR pathway (*PIK3CA, PTEN, AKT3, TSC1, TSC2, RHEB, MTOR*), and 31 samples (19 MCD, 11 LEAT, and one HS) with variants in genes of the GATOR1 complex (*DEPDC5, NPRL2*, and *NPRL3*). A detailed summary of the reclassification and genotype-phenotype analysis for the established LEAT and MCD genes is provided in the supplementary material and Figure 2C.

Major novel findings of our analysis include eight somatic *PTPN11* SNV across seven LEAT samples (three in cancer-driving sites) and eight samples with somatic duplications covering *PTPN11* (Figure 3C). Despite a clear association with LEAT, no specific LEAT histopathological subtype was strongly associated with *PTPN11* alterations; variants occurred in ganglioglioma, DNET, and three different glioma subtypes (Figure 3C, Supplementary tables 3 and 4). Second, we observed a novel association between polymicrogyria and variants in *NRAS*, with 17% (2/12) of polymicrogyria samples having an activating variant in *NRAS*. Both variants (p.Q61K and p.Q61R) have been well recognized in the literature as cancer driver variants (Figure 4A).^62,66^ Conformational changes of the *NRAS* Q61 mutated protein lead to a prolonged active state (guanosine-5’-triphosphate GTP-bound) compared to the wildtype form, with variants such as Q61R having an increased affinity for GTP, a slower rate of GTP exchange, and a lower rate of intrinsic GTP hydrolysis relative to wildtype (Figure 4B).^67^ Upon microscopic re-review, both samples with *NRAS* somatic variants had a concordant complex MCD phenotype composed of polymicrogyria and nodular heterotopia with tumor-like glioneuronal growth patterns (Figure 4C-H). Lastly, microscopic re-review of two MCD samples with duplications of the q arm of chromosome 1q21-q44 that included *AKT3* revealed a novel genotype-phenotype association. Both samples had a concordant phenotype of FCD 2a with hyaline astrocytic inclusions, a rare and seldom reported epileptogenic brain lesion (Figure 4I-K).^68^

**Figure 4:**
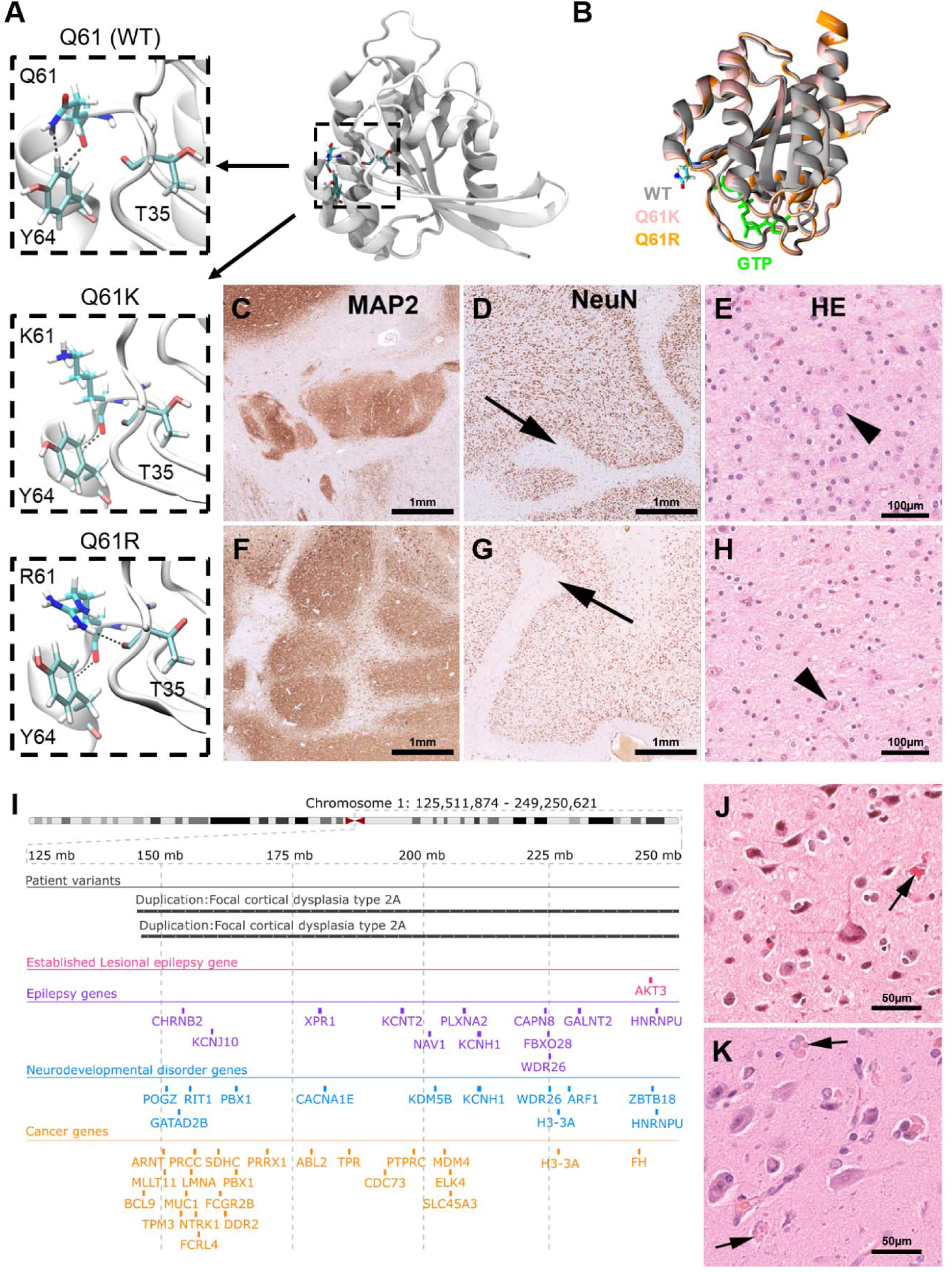
Genotype-phenotype analysis of complex malformations of cortical development with the *NRAS* mutation Q61. **(A)** Crystal structure of the wildtype (WT) N-Ras protein. Close-up of the 61^st^ amino acid site highlights changes in intermolecular interactions between different residues at position 61 and nearby positions T35 and Y64. (**B)** Overlap of the active GTP-bound 3D structures of WT, Q61K, and Q61R N-Ras highlighting changes in structure due to the introduced amino acid changes that increase the protein’s affinity to GTP. **(C-E)** A female patient in her twenties, with right temporal lobe epilepsy secondary to polymicrogyria seen on MRI. Histopathology examination of the surgical resection sample revealed a complex cortical malformation with nodular heterotopia shown in **C**, polymicrogyria (arrow in **D**), and a tumor-like glio-neuronal growth pattern resembling DNET in **E**. The arrow in **C** points to a dysplastic neuronal cell element. **(F-H)** A male patient in his twenties, with left temporal lobe epilepsy and complex malformation seen on MRI. Histopathology examination of the surgical resection sample confirmed a complex cortical malformation with nodular heterotopia shown in **F**, polymicrogyria (arrow in **G**), and a tumor-like glio-neuronal growth pattern in **H** (arrow showing a dysplastic neuronal cell). (**I-K)** Genomic coordinates of two large patient somatic duplications at 1q21-q44. Both duplications cover the established lesional epilepsy gene *AKT3*, as well as additional genes of potential interest. Breakpoints of both somatic duplications are similar. Histopathology findings were remarkably similar in both cases, showing hyaline astrocytic inclusions (arrows in **J** and **K**) next to FCD 2a. MAP2 = immunohistochemistry using antibodies directed against microtubule-associated protein 2, NeuN = immunohistochemistry using antibodies directed against the neuronal nucleus epitope, HE = hematoxylin and eosin staining.

## Discussion

We generated deep (>350x) whole-exome sequencing (WES) data to identify somatic SNV and whole-genome genotyping data to identify somatic CNV across surgically-resected epileptogenic brain lesions from 474 individuals with focal drug-resistant epilepsy. While other studies performed targeted sequencing of lesional brain tissue from <100 individuals^16^, or WES in <130 individuals^69^, our study sample represents the largest cohort of epilepsy-associated brain lesions analyzed through deep whole-exome analysis. Using this rich source of data, we demonstrated differential somatic variant profiles across LEAT, MCD, and HS. For LEAT and MCD, we confirmed previously reported and identified novel pathology-gene associations and explored the role of genetic information in clinical characteristics and histopathological classification.

We observed a differential somatic variant burden across the three most common epileptogenic pathologies (LEAT, MCD, and HS). LEAT showed a higher burden of somatic SNV and CNV than MCD or HS. This finding indicates a proliferative advantage of mutated tumor cells and conclusively shows for the first time that the genetic architecture of epilepsy-associated tumors is different from that underlying MCD or HS. We showed that increasing numbers of somatic SNV and fractions of mutated tumor cells are correlated with later age at seizure onset and older age at surgery in LEAT. This finding is in line with studies that found positive correlations between higher somatic burden and later age at diagnosis for all human genes and all cancer-associated genes across all types of cancer.^70^ We confirmed previously reported and discovered novel lesional epilepsy-associated genes using a combined statistical genetic and biological approach. Our study confirms that somatic variants affecting 12/15 recently established lesional epilepsy genes^5,14–16^ represent common genetic etiologies of lesional epilepsies (29.1%, 138/474 pathogenic variant carriers in our cohort across 12 genes). We did not observe somatic variants in *RHEB, MYB*, and *MYBL1*, in which previously identified pathogenic variants are primarily structural rearrangements such as gene fusions and CNV.^5,14^

Our study could not provide evidence for a significant role of somatic pathogenic SNV and CNV in the etiology of HS. We analyzed 97 individuals with HS, and did not identify any genetic association with HS. However, we identified a small subset of patients with somatic variants in recognized lesional epilepsy genes or the newly described *PTPN11*. Whether these variants are indeed contributing to rare subtypes of HS needs to be clarified in future studies. To the best of our knowledge, this is the first large-scale exome-level investigation of genetic variants in surgically resected HS tissues.

The majority of CNV (>69%) affected one of the 15 established lesional epilepsy genes.^5,14^ Microscopic re-review of these samples led to the identification of a novel genotype-phenotype association between duplications at q21-q44 of chromosome 1 which affects *AKT3* and many additional genes and a FCD 2a phenotype with hyaline astrocytic inclusions. This is the first report of a genotype-phenotype correlation for this lesion type and expands the spectrum of MCD phenotypes associated with somatic duplications of chromosome 1q. Compared to our study, previously reported 1q duplications were larger in size and observed in individuals with larger lesions such as polymicrogyria.^59,64,65^ Furthermore, we identified nine cases with large somatic CNN-LOH regions overlapping lesional epilepsy genes with pathogenic mechanisms known follow the “two hit” model (*TSC1, TSC2, DEPDC5, PTEN, FGFR1*, and *NF1*).^17,71^ For five of these (with the exception of *TSC1*), we identified an accompanying pathogenic germline SNV in the same gene. This result is in line with previous lesional epilepsy studies that identified CNN-LOH as a somatic second hit for germline variants in *DEPDC5, TSC1*, and *TSC2*.^17,72^ Overall, our findings add to a growing body of evidence that somatic CNV are involved in the etiology of epilepsy-associated brain lesions.^59,73^

Our exome-wide somatic variant enrichment analysis provides the first statistical evidence for *PTPN11* as a novel candidate gene for LEAT. The *PTPN11* gene encodes the Protein Tyrosine Phosphatase Non-Receptor Type 11 that regulates the RAS-RAF-MAPK signaling pathway. Heterozygous mutations in *PTPN11* cause ∼50% of all Noonan Syndrome cases^74^, a germline overgrowth disorder where multiple cases of co-occurring epileptic tumors have been reported.^52,75,76^ Interestingly, PTPN11 regulates the PI3K-AKT-MTOR and the RAS-RAF-MAPK pathway^77^ and plays a role in the development of human white matter microstructure.^53^ Furthermore, two individuals with FCD type 3 and somatic *PTPN11* mutation have been recently reported.^78^ However, five of the six carriers with LEAT and somatic *PTPN11* SNV had other potentially pathogenic somatic SNV in *TSC2, FGFR1, NF1*, or *BRAF*. This finding suggests a modifier role for *PTPN11* in LEAT, as shown for other components of the RAS-RAF-MAPK signaling pathway.^79^ Confirming the role of *PTPN11* in LEAT will require more in-depth evaluations of large LEAT cohorts as well as comprehensive animal models.^51,80^

We identified three additional candidate genes for lesional epilepsy within the RAS-RAF-MAPK pathway (i.e. *NF1, KRAS*, and *NRAS*). Our gene burden analysis identified an enrichment of somatic *NF1* variants in LEAT. Similar to *PTPN11*, no particular LEAT pathology was associated with *NF1* variants, although alterations of *NF1* have been previously identified in Ganglioglioma.^5,6^ Interestingly, we also identified somatic variants in *NF1* in four FCD 2 and one complex MCD (Figure 2C). Although HS and FCD 3a have been described in a subset of germline neurofibromatosis type 1, our study is the first direct report of *NF1* variants in *bona fide* MCD.^81,82^ We also identified an individual with sporadic meningioangiomatosis and a brain somatic cancer-driving variant in *KRAS*. Germline meningioangiomatosis is associated with neurofibromatosis type 2, but no causal gene for the sporadic form is known yet.^83^ Lastly, we identified two established cancer-driving SNV in *NRAS* in two individuals with a distinct and histopathologically-concordant complex MCD phenotype composed of polymicrogyria and nodular heterotopia with tumor-like glioneuronal growth patterns (Figure 4). The identification of potentially pharmacologically targetable *NRAS* variants^84^ in such patients may present a promising target for precision medicine when epilepsy surgery cannot achieve successful seizure control.^85^ Finally, we disclosed the genetic information for a histopathology re-review for all 151 individuals carrying somatic variants in known or novel candidate genes for lesional epilepsy. 21.2% of these cases were either reclassified *post hoc* (e.g., from mMCD to MOGHE or FCD 2a to FCD 2b) or the diagnosis was refined from the initial histopathological diagnosis (e.g. FCD2a to FCD2a with hyaline inclusions) which is in line with the results from a recent international agreement study of 22 epilepsy surgery samples.^20^

It is important to note that our exome-wide SNV and genome-wide CNV screening design casts a wide net to elucidate the genomic architecture of the lesional epilepsies and uncover potential disease-associated genes that are typically not on hypothesis-based gene panels. However, we were limited in the sequencing depth due to this approach. Our study was also limited in distinguishing between somatic variants of high allelic fraction and germline variants because of the lack of matched blood samples. Therefore, we implemented upper and lower thresholds for the allelic fraction of somatic variants that may have reduced the total number of identified somatic variants in all downstream analyses. As such, the genetic positive rate is likely higher than detected in our study and other studies with a diagnostic focus that include ultra-deep coverage sequencing and secondary validation, such as amplicon sequencing or digital PCR, should be cited to estimate diagnostic yield.

Understanding the exact molecular mechanisms involved in the etiology of epileptogenic pathologies with or without tumor activity is essential for improving treatment of drug-resistant focal epilepsy. Our study systematically shed light on the genomic landscape of the lesional epilepsies, identified four novel candidate genes for lesional epilepsy (i.e., *NRAS, KRAS, NF1*, and *PTPN11*), and observed several potentially pathogenic somatic CNV. More basic and clinical-genetic research is needed to improve surgery outcome prediction and our understanding of the etiology of the various pathologies underlining focal epilepsy.

## Supporting information

STROBE-checklist

## Data Availability

All data produced in the present study are available upon reasonable request to the authors

## Abbreviations

DNET: Dysembryoplastic neuroepithelial tumor
CNN-LOH: Copy number neutral loss of heterozygosity
CNV: Copy number variant
FCD: Focal cortical dysplasia
GG: Ganglioglioma
HME: Hemimegalencephaly
HS: Hippocampal sclerosis
LEAT: Low-grade epilepsy-associated tumor
MCD: Malformation of cortical development
mMCD: mild MCD
MOGHE: mMCD with oligodendroglial hyperplasia in epilepsy
NH: Nodular heterotopia
PMG: Polymicrogyria
PTV: Protein-truncating variant
SNV: Single nucleotide variant
VAF: Variant allele fraction
WES: Whole exome sequencing

## Acknowledgements

We wish to thank Anna Ossowski from the WGGC for her expert technical assistance.

## Supplementary material

Supplementary material is available at *Brain* online.

