## Supplementary for "The genomic landscape across 474 surgically accessible epileptogenic human brain lesions"

### Supplementary Results

The *SLC35A2* gene has been previously associated with mild MCD or FCD type 1[1,2] but was more recently confirmed as a genetic marker for the MOGHE phenotype.[3–5] We originally identified four FCD 1a or mMCD samples with *SLC35A2* variants. However, upon in-depth review of histopathological imaging, we determined that the pathological phenotypes of these samples more closely matched MOGHE and reclassified them accordingly. Thus, we detected alterations in *SLC35A2* exclusively in MOGHE samples. Accordingly, we observed a strong association between *SLC35A2* and MOGHE, with 47.1% (16/35) of MOGHE samples carrying a variant in *SLC35A2*.

As expected, alterations in *FGFR1* were exclusively identified in DNET (29%, 7/24), with multiple alterations identified in the majority of cases (71%, 5/7 *FGFR1*-DNET).[6,7] Variants in *BRAF* were strongly associated with ganglioglioma[8], with 77.6% (52/67) of detected *BRAF* alterations occurring in ganglioglioma and 57.5% (46/80) of ganglioma carrying at least one alteration in *BRAF*. Of these, the majority (40/46, 87%) carried the *BRAFV600E* variant. Furthermore, *BRAFV600E* was detected in all three pleomorphic xanthoastrocytoma[7] samples in our cohort. Of note, we observed six ganglioglioma with both a somatic *BRAF* SNV and a somatic duplication covering the *BRAF* gene (7.5%), one sample with two separate somatic duplications covering *BRAF*, and one sample with two different SNV in *BRAF* (V600E and A764V).

Our gene enrichment analysis revealed a strong association between LEAT and *PTPN11* (Figure 3). Our somatic variant gene discovery also revealed novel associations between epileptogenic brain lesions and two different RAS genes, *NRAS* and *KRAS*. Alterations in *NRAS* showed a strong association with polymicrogyria (PMG) and were exclusively detected in 17% of PMG (2/12). We detected a singular somatic *KRAS* SNV in a cancer driving site in a case of meningioangiomatosis (50%, 1/2), a lesion typically associated with the germline neurofibromatosis 2, but not somatic variants in *NF2*.[9,10] To the best of our knowledge, we are the first to report an association between RAS genes and epileptogenic lesions. Lastly, we identified an association between somatic alterations in *NF1* and epileptogenic brain lesions. We detected nine *NF1* alterations across seven LEAT, which included two ganglioglioma, two DNET, and three gliomas (Fig 3A). Among these, we identified DNET sample carrying two distinct somatic *NF1* SNV (a splicing variant and a stopgain) and a pilocytic astrocytoma carrying a somatic *NF1* PTV and a somatic CNN-LOH covering *NF1*. We also identified an additional seven *NF1* alterations across five MCD, which included four FCD II and a single PMG (Fig 3A). From these, we only detected more than one variant in the PMG, which carried three distinct *NF1* alterations (T30K, G26X, and a somatic CNN-LOH covering *NF1*). Our results expand the spectrum of epileptogenic brain lesions associated to variants in the canonical RAS-RAF-MAPK pathway and serve as a foundation for untangling the role of this pathway in lesional epilepsies.

**Supplementary Table 1: Top hits from gene enrichment analysis**

| Gene | Synonymous variants | Missense variants | Nonsense variants | Splice variants | Insertions/ deletions | $P_{global}$ (unadjusted) | $Q_{global}$ |
|---|---|---|---|---|---|---|---|
| **Hippocampal Sclerosis (n=97)** | | | | | | | |
| *NUDT14* | 0 | 0 | 0 | 0 | 1 | 0.001052 | 1 |
| *TEX45* | 0 | 0 | 0 | 0 | 1 | 0.001724 | 1 |
| *FKBP10* | 0 | 0 | 0 | 0 | 1 | 0.001858 | 1 |
| *ARNT* | 0 | 0 | 0 | 0 | 1 | 0.00216 | 1 |
| *UNC5C* | 0 | 0 | 0 | 0 | 1 | 0.002331 | 1 |
| **Malformations of cortical development (n=223)** | | | | | | | |
| *SLC35A2* | 0 | 7 | 2 | 0 | 3 | 0 | **0** |
| *MTOR* | 0 | 11 | 0 | 0 | 1 | 2.56E-10 | **2.57E-06** |
| *AKT3* | 0 | 3 | 0 | 0 | 0 | 3.47E-04 | 1 |
| *OR9G1* | 0 | 4 | 0 | 0 | 0 | 7.59E-04 | 1 |
| *OR9G9* | 0 | 4 | 0 | 0 | 0 | 7.59E-04 | 1 |
| *NRAS* | 0 | 2 | 0 | 0 | 0 | 0.002669 | 1 |
| *CD3G* | 0 | 0 | 0 | 0 | 1 | 0.003661 | 1 |
| **Low-grade epilepsy-associated tumors (n=154)** | | | | | | | |
| *BRAF* | 0 | 39 | 0 | 0 | 3 | 0 | **0** |
| *PTPN11* | 0 | 6 | 0 | 0 | 0 | 4.19E-08 | **4.21E-04** |
| *FGFR1* | 0 | 4 | 0 | 0 | 0 | 2.62E-05 | 0.175515 |
| *OR4C3* | 0 | 4 | 0 | 0 | 0 | 3.16E-04 | 1 |
| *AURKC* | 0 | 1 | 0 | 0 | 1 | 3.59E-04 | 1 |
| *NF1* | 0 | 0 | 0 | 0 | 2 | 7.76E-04 | 1 |
| *RARA* | 0 | 1 | 0 | 0 | 1 | 0.001494 | 1 |
| *HSPA2* | 0 | 1 | 0 | 0 | 1 | 0.002737 | 1 |
| *GNRH2* | 0 | 0 | 0 | 0 | 1 | 0.004502 | 1 |

**Supplementary Table 2: Identified cancer drivers**

| Gene | Known driver site | Samples with variants | Associated pathology | Specific sample pathologies |
|---|---|---|---|---|
| *BRAF* | p.V600 | 45 | LEAT | Ganglioglioma, PXA, MVNT |
| *MTOR* | p.S2215 | 3 | MCD | FCD type IIb, HME |
| | p.T1977 | 1 | MCD | FCD type IIb |
| | p.L1460 | 1 | MCD | FCD type IIb |
| *NRAS* | p.Q61 | 2 | MCD | PMG |
| *PTPN11* | p.A72 | 2 | LEAT | DNET |
| | p.E76 | 1 | LEAT | Ganglioglioma |
| *KRAS* | p.G12 | 1 | LEAT | Meningioangiomatosis |
| *PIK3CA* | p.V344 | 1 | LEAT | DNET |
| | p.E545 | 1 | MCD | HME |

**Legend:** PXA = Pleomorphic xanthoastrocytoma, MVNT = Multinodular vacuolated neuronal tumor, FCD = Focal cortical dysplasia, HME = Hemimegalencephaly, PMG = Polymicrogyria, DNET = Dysembryoplastic Neuroepithelial tumor

**Supplementary table 3: Phenotypic review of genetic-positive samples**

| Sample name | Lesion category | Original pathology | Genetic diagnosis | Revised pathology | Additional information | Conclusion after histopathological review |
|---|---|---|---|---|---|---|
| MCD_EEBB_26 | MCD | HME | *PIK3CA* | Complex MCD | HME / PMG / FCD 2a | phenotype refinement |
| MCD_EEBB_57 | MCD | HME | *AKT3, MTOR* | Complex MCD | HME / PMG / FCD 2a | phenotype refinement |
| MCD_EEBB_52 | MCD | HME | *PIK3CA* | Complex MCD | HME / PMG / FCD 2a | phenotype refinement |
| MCD_EEBB_10 | MCD | PMG | *NRAS* | Complex MCD | PMG / NH / FCD 2a / GG | phenotype refinement |
| MCD_EEBB_89 | MCD | PMG | *NF1* SNV + CNN-LOH | Complex MCD | PMG / FCD 1b | phenotype refinement |
| MCD_EEBB_65 | MCD | PMG | *NRAS* | Complex MCD | PMG / NH / FCD 2a / DNET | phenotype refinement |
| MCD_EEBB_21 | MCD | PMG | *PIK3CA* | Complex MCD | HME / PMG / FCD 2a | reclassification |
| MCD_CCF_93 | MCD | FCD (NOS) | *AKT3* | Complex MCD | HME / PMG / FCD 2a | reclassification |
| MCD_CCF_94 | MCD | FCD (NOS) | *PTEN* SNV + CNN-LOH | Complex MCD | HME / PMG / FCD 2a / FCD 1a | reclassification |
| MCD_CCF_110 | MCD | FCD (NOS) | *AKT3* | Complex MCD | HME / PMG / FCD 2a | reclassification |
| MCD_CCF_32 | MCD | FCD (NOS) | *NPRL3* | Complex MCD | PMG / FCD 2a / FCD 1a | reclassification |
| MCD_CCF_104 | MCD | FCD (NOS) | *MTOR, TSC2* SNV + CNN-LOH | Complex MCD | PMG / FCD 2b | reclassification |
| MCD_CCF_92 | MCD | FCD (NOS) | *MTOR* | FCD 2b | | reclassification |
| MCD_CCF_101 | MCD | FCD (NOS) | *MTOR* | FCD 2b | | reclassification |
| MCD_CCF_29 | MCD | FCD (NOS) | *TSC2* | Cortical tuber | FCD 2b | reclassification |
| MCD_CCF_55 | MCD | FCD (NOS) | *TSC2* | Cortical tuber | FCD 2b | reclassification |
| MCD_CCF_56 | MCD | FCD (NOS) | *TSC2* | Cortical tuber | FCD 2b | reclassification |
| MCD_CCF_27 | MCD | FCD 2b | *TSC2* | Cortical tuber | FCD 2b | reclassification |
| MCD_EEBB_76 | MCD | FCD 2b | *MTOR* | Complex MCD | PMG / FCD 2b | reclassification |
| MCD_EEBB_68 | MCD | FCD 2b | *DEPDC5* CNN-LOH | FCD 2a | | reclassification |
| MCD_CCF_62 | MCD | FCD 2a | Chr1q gain | FCD 2a | Hyaline astrocytic inclusions | phenotype refinement |
| MCD_EEBB_110 | MCD | MOGHE | Chr1q gain | FCD 2a | Hyaline astrocytic inclusions | reclassification |
| MCD_CCF_30 | MCD | mMCD | *DEPDC5* | FCD 2a | | reclassification |
| MCD_CCF_80 | MCD | mMCD | *DEPDC5* | FCD 2a | | reclassification |
| MCD_EEBB_100 | MCD | mMCD | *SLC35A2* | MOGHE | | reclassification |
| MCD_CCF_79 | MCD | FCD 1a | *SLC35A2* | MOGHE | | reclassification |
| MCD_EEBB_58 | MCD | FCD 1a | *SLC35A2* | MOGHE | | reclassification |
| MCD_EEBB_87 | MCD | FCD 1a | *SLC35A2* | MOGHE | | reclassification |
| MCD_CCF_77 | MCD | FCD 1a | *DEPDC5* | MOGHE | FCD 1a | reclassification |
| LEAT_CCF_6 | LEAT | LEAT (NOS) | *PTPN11* | DNET | | reclassification |
| LEAT_CCF_37 | LEAT | LEAT (NOS) | Chr7 gain, *TSC2* (somatic) SNV + CNN-LOH | Subependymal giant cell astrocytoma | | reclassification |

Legend to supplementary table 3: MCD = Malformation of cortical development, LEAT = Low-grade epilepsy-associated tumor, FCD = Focal cortical dysplasia, NOS = Not otherwise specified, HME = Hemimegalencephaly, PMG = Polymicrogyria, mMCD = mild MCD, MOGHE = mMCD with oligodendroglial hyperplasia in epilepsy, NH = nodular heterotopia, DNET = Dysembryoplastic neuroepithelial tumor, IDA = Isomorphic diffuse astrocytoma, PXA = Pleomorphic xanthoastrocytoma, MVNT = multinodular vacuolated neuronal tumor, SEGA = Subependymal giant cell astrocytoma, GG = Ganglioglioma, CNN-LOH = Copy number neutral loss of heterozygosity

**Supplementary Table 4: Reported SNV in 19 lesional epilepsy genes**

| Sample | Sex | Age at onset | Age at surgery | Lesion group | Pathology | Additional findings | Gene | Variant position(hg19) | Transcript ID | cDNA change | Amino acid change | Variant class | Allelic fraction |
|---|---|---|---|---|---|---|---|---|---|---|---|---|---|
| HS_EEBB_20 | F | 0-4 | 45-49 | HS | HS Type 1 | FCD 3a | *MTOR* | chr1:11188137 | NM_004958 | c.C5957G | p.A1986G | Missense | 0.47 |
| HS_EEBB_41 | F | 0-4 | 35-39 | HS | HS Type 1 | | *PTPN11* | chr12:112926885 | NM_002834 | c.C1505T | p.S502L | Missense | 0.02 |
| HS_EEBB_53 | M | 0-4 | 45-49 | HS | HS Type 1 | | *TSC1* | chr9:135772010 | NM_000368 | c.G3107A | p.G1036E | Missense | 0.50 |
| HS_EEBB_71 | M | 45-49 | 70-74 | HS | HS Type 1 | | *MTOR* | chr1:11301631 | NM_004958 | c.C1520T | p.P507L | Missense | 0.47 |
| HS_EEBB_17 | M | 20-24 | 25-29 | HS | HS Type 2 | FCD 3a | *PTEN* | chr10:89720649 | NM_001304717 | c.1321-2A>T | Splicing | PTV | 0.03 |
| LEAT_EEBB_1 | F | 10-14 | 15-19 | LEAT | Ganglioglioma | | *BRAF* | chr7:140453136 | NM_004333 | c.T1799A | p.V600E | Missense | 0.17 |
| LEAT_EEBB_10 | F | 5-9 | 15-19 | LEAT | Ganglioglioma | | *BRAF* | chr7:140453136 | NM_004333 | c.T1799A | p.V600E | Missense | 0.15 |
| LEAT_EEBB_10 | F | 5-9 | 15-19 | LEAT | Ganglioglioma | | *PTPN11* | chr12:112888210 | NM_002834 | c.G226A | p.E76K | Missense | 0.02 |
| LEAT_EEBB_100 | M | 10-14 | 15-19 | LEAT | Ganglioglioma | | *BRAF* | chr7:140453136 | NM_004333 | c.T1799A | p.V600E | Missense | 0.17 |
| LEAT_EEBB_100 | M | 10-14 | 15-19 | LEAT | Ganglioglioma | | *BRAF* | chr7:140426307 | NM_001354609 | c.C2291T | p.A764V | Missense | 0.03 |
| LEAT_EEBB_103 | F | 15-19 | 20-24 | LEAT | Ganglioglioma | HS Type 1 | *BRAF* | chr7:140453136 | NM_004333 | c.T1799A | p.V600E | Missense | 0.36 |
| LEAT_EEBB_106 | F | 0-4 | 30-34 | LEAT | Ganglioglioma | HS Type 1 | *BRAF* | chr7:140453136 | NM_004333 | c.T1799A | p.V600E | Missense | 0.04 |
| LEAT_EEBB_107 | M | 0-4 | 5-9 | LEAT | Ganglioglioma | | *BRAF* | chr7:140453136 | NM_004333 | c.T1799A | p.V600E | Missense | 0.04 |
| LEAT_EEBB_108 | F | 5-9 | 5-9 | LEAT | Ganglioglioma | | *BRAF* | chr7:140453136 | NM_004333 | c.T1799A | p.V600E | Missense | 0.05 |
| LEAT_EEBB_12 | M | 0-4 | 0-4 | LEAT | Ganglioglioma | | *BRAF* | chr7:140453136 | NM_004333 | c.T1799A | p.V600E | Missense | 0.10 |
| LEAT_EEBB_14 | M | | 15-19 | LEAT | Ganglioglioma | | *BRAF* | chr7:140453136 | NM_004333 | c.T1799A | p.V600E | Missense | 0.04 |
| LEAT_EEBB_15 | M | 15-19 | 15-19 | LEAT | Ganglioglioma | HS | *BRAF* | chr7:140453136 | NM_004333 | c.T1799A | p.V600E | Missense | 0.13 |
| LEAT_EEBB_15 | M | 15-19 | 15-19 | LEAT | Ganglioglioma | HS | *PTPN11* | chr12:112888165 | NM_002834 | c.G181A | p.D61N | Missense | 0.03 |
| LEAT_EEBB_15 | M | 15-19 | 15-19 | LEAT | Ganglioglioma | HS | *PTPN11* | chr12:112891083 | NM_002834 | c.G417C | p.E139D | Missense | 0.03 |
| LEAT_EEBB_22 | M | | 0-4 | LEAT | Ganglioglioma | HS Type 1 | *BRAF* | chr7:140453136 | NM_004333 | c.T1799A | p.V600E | Missense | 0.09 |
| LEAT_EEBB_22 | M | | 0-4 | LEAT | Ganglioglioma | HS Type 1 | *NF1* | chr17:29579966 | NM_001042492 | c.A4121G | p.H1374R | Missense | 0.44 |
| LEAT_EEBB_22 | M | | 0-4 | LEAT | Ganglioglioma | HS Type 1 | *TSC2* | chr16:2130303 | NM_000548 | c.G3535A | p.E1179K | Missense | 0.44 |
| LEAT_EEBB_27 | M | 15-19 | 15-19 | LEAT | Ganglioglioma | | *BRAF* | chr7:140453136 | NM_004333 | c.T1799A | p.V600E | Missense | 0.15 |
| LEAT_EEBB_32 | F | 20-24 | 20-24 | LEAT | Ganglioglioma | | *BRAF* | chr7:140453137 | NM_004333 | c.1798delinsACAG | p.T599_V600insT | Missense | 0.26 |
| LEAT_EEBB_34 | M | | 0-4 | LEAT | Ganglioglioma | | *BRAF* | chr7:140453136 | NM_004333 | c.T1799A | p.V600E | Missense | 0.19 |
| LEAT_EEBB_37 | M | 0-4 | 40-44 | LEAT | Ganglioglioma | HS Type 1 | *BRAF* | chr7:140453136 | NM_004333 | c.T1799A | p.V600E | Missense | 0.04 |
| LEAT_EEBB_39 | F | 0-4 | 25-29 | LEAT | Ganglioglioma | | *BRAF* | chr7:140453136 | NM_004333 | c.T1799A | p.V600E | Missense | 0.12 |

| | | | | | | | | | | | | | |
|---|---|---|---|---|---|---|---|---|---|---|---|---|---|
| LEAT_EEBB_40 | F | 10-14 | 25-29 | LEAT | Ganglioglioma | HS Type 1 | *BRAF* | chr7:140453136 | NM_004333 | c.T1799A | p.V600E | Missense | 0.20 |
| LEAT_EEBB_42 | F | 20-24 | 30-34 | LEAT | Ganglioglioma | | *BRAF* | chr7:140453136 | NM_004333 | c.T1799A | p.V600E | Missense | 0.08 |
| LEAT_EEBB_43 | F | 0-4 | 0-4 | LEAT | Ganglioglioma | | *BRAF* | chr7:140453136 | NM_004333 | c.T1799A | p.V600E | Missense | 0.25 |
| LEAT_EEBB_44 | M | 0-4 | 0-4 | LEAT | Ganglioglioma | | *BRAF* | chr7:140453136 | NM_004333 | c.T1799A | p.V600E | Missense | 0.04 |
| LEAT_EEBB_47 | F | 0-4 | 0-4 | LEAT | Ganglioglioma | | *BRAF* | chr7:140453136 | NM_004333 | c.T1799A | p.V600E | Missense | 0.08 |
| LEAT_EEBB_52 | M | 10-14 | 20-24 | LEAT | Ganglioglioma | | *BRAF* | chr7:140453136 | NM_004333 | c.T1799A | p.V600E | Missense | 0.09 |
| LEAT_EEBB_53 | F | 10-14 | 15-19 | LEAT | Ganglioglioma | | *BRAF* | chr7:140453136 | NM_004333 | c.T1799A | p.V600E | Missense | 0.04 |
| LEAT_EEBB_54 | M | 0-4 | 5-9 | LEAT | Ganglioglioma | | *BRAF* | chr7:140453136 | NM_004333 | c.T1799A | p.V600E | Missense | 0.18 |
| LEAT_EEBB_57 | M | 0-4 | 5-9 | LEAT | Ganglioglioma | | *BRAF* | chr7:140453136 | NM_004333 | c.T1799A | p.V600E | Missense | 0.13 |
| LEAT_EEBB_61 | M | 0-4 | 0-4 | LEAT | Ganglioglioma | | *BRAF* | chr7:140453136 | NM_004333 | c.T1799A | p.V600E | Missense | 0.15 |
| LEAT_EEBB_62 | M | 45-49 | 45-49 | LEAT | Ganglioglioma | | *TSC1* | chr9:135772994 | NM_000368 | c.G2629T | p.V877L | Missense | 0.03 |
| LEAT_EEBB_63 | M | 10-14 | 15-19 | LEAT | Ganglioglioma | | *BRAF* | chr7:140453136 | NM_004333 | c.T1799A | p.V600E | Missense | 0.12 |
| LEAT_EEBB_65 | M | 0-4 | 5-9 | LEAT | Ganglioglioma | | *BRAF* | chr7:140453136 | NM_004333 | c.T1799A | p.V600E | Missense | 0.23 |
| LEAT_EEBB_67 | M | | 5-9 | LEAT | Ganglioglioma | | *BRAF* | chr7:140453136 | NM_004333 | c.T1799A | p.V600E | Missense | 0.12 |
| LEAT_EEBB_73 | M | 0-4 | 5-9 | LEAT | Ganglioglioma | | *BRAF* | chr7:140453136 | NM_004333 | c.T1799A | p.V600E | Missense | 0.14 |
| LEAT_EEBB_74 | M | 5-9 | 10-14 | LEAT | Ganglioglioma | | *BRAF* | chr7:140453136 | NM_004333 | c.T1799A | p.V600E | Missense | 0.14 |
| LEAT_EEBB_76 | F | 5-9 | 15-19 | LEAT | Ganglioglioma | | *BRAF* | chr7:140453136 | NM_004333 | c.T1799A | p.V600E | Missense | 0.16 |
| LEAT_EEBB_78 | M | 0-4 | 5-9 | LEAT | Ganglioglioma | HS Type 1 | *BRAF* | chr7:140453136 | NM_004333 | c.T1799A | p.V600E | Missense | 0.04 |
| LEAT_EEBB_80 | M | 5-9 | 5-9 | LEAT | Ganglioglioma | | *BRAF* | chr7:140453136 | NM_004333 | c.T1799A | p.V600E | Missense | 0.19 |
| LEAT_EEBB_84 | M | 0-4 | 0-4 | LEAT | Ganglioglioma | HS Type 2 | *BRAF* | chr7:140453136 | NM_004333 | c.T1799A | p.V600E | Missense | 0.21 |
| LEAT_EEBB_84 | M | 0-4 | 0-4 | LEAT | Ganglioglioma | HS Type 2 | *PIK3CA* | chr3:178942554 | NM_006218 | c.C2361A | p.D787E | Missense | 0.47 |
| LEAT_EEBB_85 | M | | 30-34 | LEAT | Ganglioglioma | | *BRAF* | chr7:140453136 | NM_004333 | c.T1799A | p.V600E | Missense | 0.13 |
| LEAT_EEBB_90 | F | | 45-49 | LEAT | Ganglioglioma | | *BRAF* | chr7:140453136 | NM_004333 | c.T1799A | p.V600E | Missense | 0.06 |
| LEAT_EEBB_90 | F | | 45-49 | LEAT | Ganglioglioma | | *PTEN* | chr10:89720649 | NM_001304717 | c.1321-2A>T | Splicing | PTV | 0.06 |
| LEAT_EEBB_91 | F | 5-9 | 10-14 | LEAT | Ganglioglioma | | *BRAF* | chr7:140477841 | NM_004333 | c.1465_1467delinsA | p.A489Tfs*12 | PTV | 0.03 |
| LEAT_EEBB_94 | M | 5-9 | 5-9 | LEAT | Ganglioglioma | | *BRAF* | chr7:140453136 | NM_004333 | c.T1799A | p.V600E | Missense | 0.41 |
| LEAT_EEBB_96 | M | 5-9 | 5-9 | LEAT | Ganglioglioma | HS Type 1 | *BRAF* | chr7:140453136 | NM_004333 | c.T1799A | p.V600E | Missense | 0.11 |
| LEAT_EEBB_98 | M | 15-19 | 15-19 | LEAT | Ganglioglioma | | *BRAF* | chr7:140453136 | NM_004333 | c.T1799A | p.V600E | Missense | 0.12 |
| LEAT_EEBB_99 | F | 0-4 | 5-9 | LEAT | Ganglioglioma | | *BRAF* | chr7:140453136 | NM_004333 | c.T1799A | p.V600E | Missense | 0.12 |
| LEAT_CCF_6 | F | 30-34 | 30-34 | LEAT | DNET | | *PTPN11* | chr12:112926900 | NM_002834 | c.C1520A | p.T507K | Missense | 0.02 |

| | | | | | | | | | | | | | |
|---|---|---|---|---|---|---|---|---|---|---|---|---|---|
| LEAT_EEBB_105 | F | 35-39 | 35-39 | LEAT | DNET | | *FGFR1* | chr8:38272308 | NM_001174067 | c.A2059G | p.K687E | Missense | 0.18 |
| LEAT_EEBB_105 | F | 35-39 | 35-39 | LEAT | DNET | | *FGFR1* | chr8:38274850 | NM_001174067 | c.A1730G | p.N577S | Missense | 0.11 |
| LEAT_EEBB_105 | F | 35-39 | 35-39 | LEAT | DNET | | *NF1* | chr17:29554604 | NM_001042492 | c.2389delG | p.A797Pfs*24 | PTV | 0.04 |
| LEAT_EEBB_33 | M | 10-14 | 15-19 | LEAT | DNET | | *PIK3CA* | chr3:178952085 | NM_006218 | c.A3140G | p.H1047R | Missense | 0.03 |
| LEAT_EEBB_49 | F | 30-34 | 30-34 | LEAT | DNET | | *FGFR1* | chr8:38274849 | NM_001174067 | c.C1731A | p.N577K | Missense | 0.18 |
| LEAT_EEBB_49 | F | 30-34 | 30-34 | LEAT | DNET | | *NF1* | chr17:29665035 | NM_001042492 | c.6705-8_6705-1delinsA | Splicing | PTV | 0.05 |
| LEAT_EEBB_49 | F | 30-34 | 30-34 | LEAT | DNET | | *NF1* | chr17:29552151 | NM_001042492 | c.C1884G | p.Y628X | PTV | 0.05 |
| LEAT_EEBB_49 | F | 30-34 | 30-34 | LEAT | DNET | | *PTPN11* | chr12:112888199 | NM_002834 | c.C215T | p.A72V | Missense | 0.03 |
| LEAT_EEBB_6 | M | 10-14 | 10-14 | LEAT | DNET | | *FGFR1* | chr8:38272308 | NM_001174067 | c.A2059G | p.K687E | Missense | 0.42 |
| LEAT_EEBB_6 | M | 10-14 | 10-14 | LEAT | DNET | | *FGFR1* | chr8:38272308 | NM_001174067 | c.A2059C | p.K687Q | Missense | 0.42 |
| LEAT_EEBB_6 | M | 10-14 | 10-14 | LEAT | DNET | | *FGFR1* | chr8:38272319 | NM_001174067 | c.A2048G | p.D683G | Missense | 0.41 |
| LEAT_EEBB_60 | M | 15-19 | 20-24 | LEAT | DNET | | *FGFR1* | chr8:38272308 | NM_001174067 | c.A2059G | p.K687E | Missense | 0.04 |
| LEAT_EEBB_60 | M | 15-19 | 20-24 | LEAT | DNET | | *FGFR1* | chr8:38272322 | NM_001174067 | c.T2045C | p.I682T | Missense | 0.03 |
| LEAT_EEBB_81 | M | 45-49 | 55-59 | LEAT | DNET | | *FGFR1* | chr8:38274849 | NM_001174067 | c.C1731A | p.N577K | Missense | 0.23 |
| LEAT_EEBB_81 | M | 45-49 | 55-59 | LEAT | DNET | | *PTPN11* | chr12:112888199 | NM_002834 | c.C215G | p.A72G | Missense | 0.02 |
| LEAT_EEBB_83 | M | 5-9 | 10-14 | LEAT | DNET | | *FGFR1* | chr8:38272302 | NM_001174067 | c.A2065C | p.T689P | Missense | 0.31 |
| LEAT_EEBB_83 | M | 5-9 | 10-14 | LEAT | DNET | | *FGFR1* | chr8:38272307 | NM_001174067 | c.A2060T | p.K687M | Missense | 0.30 |
| LEAT_EEBB_83 | M | 5-9 | 10-14 | LEAT | DNET | | *FGFR1* | chr8:38272321 | NM_001174067 | c.C2046G | p.I682M | Missense | 0.30 |
| LEAT_EEBB_83 | M | 5-9 | 10-14 | LEAT | DNET | | *PIK3CA* | chr3:178921549 | NM_006218 | c.T1031G | p.V344G | Missense | 0.03 |
| LEAT_EEBB_2 | F | | 35-39 | LEAT | Angiocentric glioma | | *DEPDC5* | chr22:32206593 | NM_001242896 | c.G1411A | p.G471S | Missense | 0.54 |
| LEAT_EEBB_31 | M | 10-14 | 10-14 | LEAT | MA | FCD 3c | *KRAS* | chr12:25398284 | NM_033360 | c.G35A | p.G12D | Missense | 0.05 |
| LEAT_EEBB_113 | M | 0-4 | 44-49 | LEAT | MVNT | | *BRAF* | chr7:140453138 | NM_004333 | c.1797delinsTACA | p.T599_V600insT | Missense | 0.14 |
| LEAT_EEBB_95 | F | 5-9 | 35-39 | LEAT | Pilocytic Astrocytoma | | *NF1* | chr17:29677325 | NM_001042492 | c.7446_7450delinsC | p.S2484Ifs*4 | PTV | 0.13 |
| LEAT_EEBB_38 | M | 10-14 | 15-19 | LEAT | PXA | | *BRAF* | chr7:140453136 | NM_004333 | c.T1799A | p.V600E | Missense | 0.12 |
| LEAT_EEBB_56 | M | 10-14 | 15-19 | LEAT | PXA | | *BRAF* | chr7:140453136 | NM_004333 | c.T1799A | p.V600E | Missense | 0.41 |
| LEAT_EEBB_56 | M | 10-14 | 15-19 | LEAT | PXA | | *NF1* | chr17:29559097 | NM_001042492 | c.G3204T | p.L1068F | Missense | 0.28 |
| LEAT_EEBB_59 | M | | 15-19 | LEAT | PXA | | *BRAF* | chr7:140453136 | NM_004333 | c.T1799A | p.V600E | Missense | 0.22 |
| LEAT_CCF_37 | M | 0-4 | 5-9 | LEAT | TSC | SEGA | *TSC2* | chr16:2106223 | NM_000548 | c.626_628delinsC | p.A210Vfs*24 | PTV | 0.02 |
| LEAT_EEBB_17 | F | 0-4 | 5-9 | LEAT | TSC | SEGA | *PTPN11* | chr12:112888165 | NM_002834 | c.G181C | p.D61H | Missense | 0.05 |

| | | | | | | | | | | | | | |
|---|---|---|---|---|---|---|---|---|---|---|---|---|---|
| LEAT_EEBB_17 | F | 0-4 | 5-9 | LEAT | TSC | SEGA | *TSC2* | chr16:2106719 | NM_000548 | c.724dupA | p.T242Nfs*96 | PTV | 0.46 |
| MCD_CCF_19 | M | 10-14 | 25-29 | MCD | MOGHE | | *SLC35A2* | chrX:48762551 | NM_005660 | c.632_635del | p.L211Pfs*137 | PTV | 0.05 |
| MCD_CCF_43 | M | 0-4 | 5-9 | MCD | MOGHE | | *SLC35A2* | chrX:48762557 | NM_005660 | c.G629A | p.C210Y | Missense | 0.05 |
| MCD_CCF_77 | F | 0-4 | 0-4 | MCD | MOGHE | FCD1a | *DEPDC5* | chr22:32161000 | NM_001242896 | c.G233C | p.R78P | Missense | 0.40 |
| MCD_CCF_79 | F | 0-4 | 0-4 | MCD | MOGHE | | *SLC35A2* | chrX:48763748 | NM_005660 | c.C347A | p.A116E | Missense | 0.14 |
| MCD_CCF_108 | M | 5-9 | 25-29 | MCD | MOGHE | | *SLC35A2* | chrX:48767173 | NM_005660 | c.192delinsACCGC | p.F65Pfs*30 | PTV | 0.24 |
| MCD_EEBB_2 | F | 0-4 | 5-9 | MCD | MOGHE | | *SLC35A2* | chrX:48762251 | NM_005660 | c.C935T | p.S312F | Missense | 0.08 |
| MCD_EEBB_17 | F | 20-24 | 55-59 | MCD | MOGHE | | *SLC35A2* | chrX:48762671 | NM_005660 | c.T515C | p.L172P | Missense | 0.03 |
| MCD_EEBB_34 | F | 10-14 | 40-44 | MCD | MOGHE | | *SLC35A2* | chrX:48763729 | NM_005660 | c.366delinsTCTC | p.Y122_T123insL | Missense | 0.09 |
| MCD_EEBB_34 | F | 10-14 | 40-44 | MCD | MOGHE | | *SLC35A2* | chrX:48763730 | NM_005660 | c.364_365insC | p.Y122Sfs*6 | PTV | 0.15 |
| MCD_EEBB_37 | M | 0-4 | 25-29 | MCD | MOGHE | | *SLC35A2* | chrX:48762518 | NM_005660 | c.665_668delinsT | p.K222del | Missense | 0.29 |
| MCD_EEBB_58 | M | 0-4 | 0-4 | MCD | MOGHE | | *SLC35A2* | chrX:48762239 | NM_005660 | c.921_947delinsT | p.S308Wfs*106 | PTV | 0.11 |
| MCD_EEBB_63 | M | 0-4 | 5-9 | MCD | MOGHE | | *SLC35A2* | chrX:48762510 | NM_005660 | c.675dupA | p.G226Rfs*29 | PTV | 0.40 |
| MCD_EEBB_69 | F | 15-19 | 40-44 | MCD | MOGHE | | *SLC35A2* | chrX:48762546 | NM_005660 | c.G640C | p.G214R | Missense | 0.08 |
| MCD_EEBB_87 | F | 0-4 | 5-9 | MCD | MOGHE | | *SLC35A2* | chrX:48763773 | NM_005660 | c.C322T | p.Q108X | PTV | 0.09 |
| MCD_EEBB_97 | F | 0-4 | 15-19 | MCD | MOGHE | | *NPRL2* | chr3:50385595 | NM_006545 | c.A892C | p.I298L | Missense | 0.04 |
| MCD_EEBB_97 | F | 0-4 | 15-19 | MCD | MOGHE | | *SLC35A2* | chrX:48762281 | NM_005660 | c.C905T | p.S302F | Missense | 0.08 |
| MCD_EEBB_100 | M | | 5-9 | MCD | MOGHE | | *SLC35A2* | chrX:48763773 | NM_005660 | c.C322T | p.Q108X | PTV | 0.25 |
| MCD_EEBB_102 | F | 0-4 | 0-4 | MCD | MOGHE | | *SLC35A2* | chrX:48762251 | NM_005660 | c.C935T | p.S312F | Missense | 0.12 |
| MCD_EEBB_106 | M | 0-4 | 5-9 | MCD | MOGHE | | *SLC35A2* | chrX:48762354 | NM_005660 | c.C832T | p.Q278X | PTV | 0.48 |
| MCD_EEBB_22 | F | 5-9 | 25-29 | MCD | FCD 1a | | *MTOR* | chr1:11307914 | NM_004958 | c.C1078T | p.R360W | Missense | 0.04 |
| MCD_CCF_8 | M | 5-9 | 20-24 | MCD | FCD 2a | | *NF1* | chr17:29559092 | NM_001042492 | c.G3199T | p.D1067Y | Missense | 0.25 |
| MCD_CCF_13 | M | 0-4 | 20-24 | MCD | FCD 2a | | *NF1* | chr17:29654761 | NM_001042492 | c.C5513G | p.S1838C | Missense | 0.43 |
| MCD_CCF_13 | M | 0-4 | 20-24 | MCD | FCD 2a | | *NPRL2* | chr3:50386328 | NM_006545 | c.C562T | p.Q188X | PTV | 0.53 |
| MCD_CCF_30 | M | 0-4 | 15-19 | MCD | FCD 2a | | *DEPDC5* | chr22:32193632 | NM_001242896 | c.G814T | p.V272L | Missense | 0.46 |
| MCD_CCF_46 | F | 5-9 | 25-29 | MCD | FCD 2a | | *NPRL3* | chr16:142605 | ENST00000399953 | c.1149dupC | p.A384Rfs*71 | PTV | 0.50 |
| MCD_CCF_58 | F | 0-4 | 0-4 | MCD | FCD 2a | | *NPRL3* | chr16:143291 | ENST00000399953 | c.G957A | p.W319X | PTV | 0.56 |
| MCD_CCF_60 | F | 0-4 | 0-4 | MCD | FCD 2a | | *NPRL2* | chr3:50388010 | NM_006545 | c.73dupT | p.Y25Lfs*5 | PTV | 0.57 |
| MCD_CCF_80 | F | 45-49 | 55-59 | MCD | FCD 2a | | *DEPDC5* | chr22:32233122 | NM_001242896 | c.C2335G | p.P779A | Missense | 0.49 |

| | | | | | | | | | | | | | |
|---|---|---|---|---|---|---|---|---|---|---|---|---|---|
| MCD_CCF_83 | F | 0-4 | 5-9 | MCD | FCD 2a | | *NPRL3* | chr16:136827 | ENST00000399953 | c.1584delT | p.M529Cfs*23 | PTV | 0.48 |
| MCD_EEBB_18 | M | 5-9 | 10-14 | MCD | FCD 2a | | *BRAF* | chr7:140549970 | NM_004333 | c.A181G | p.I61V | Missense | 0.42 |
| MCD_EEBB_56 | M | 0-4 | 0-4 | MCD | FCD 2a | | *NPRL3* | chr16:143268 | ENST00000399953 | c.C980T | p.P327L | Missense | 0.42 |
| MCD_EEBB_75 | M | 0-4 | 10-14 | MCD | FCD 2a | | *DEPDC5* | chr22:32239185 | NM_001242896 | c.C2620T | p.R874X | PTV | 0.43 |
| MCD_EEBB_86 | M | 15-19 | 40-44 | MCD | FCD 2a | | *MTOR* | chr1:11217230 | NM_004958 | c.G4448A | p.C1483Y | Missense | 0.05 |
| MCD_EEBB_88 | M | 0-4 | 5-9 | MCD | FCD 2a | | *NPRL3* | chr16:143268 | ENST00000399953 | c.C980T | p.P327L | Missense | 0.46 |
| MCD_EEBB_91 | M | 5-9 | 35-39 | MCD | FCD 2a | | *DEPDC5* | chr22:32188751 | NM_001242896 | c.C715T | p.R239X | PTV | 0.42 |
| MCD_CCF_28 | F | 0-4 | 5-9 | MCD | FCD 2b | | *TSC2* | chr16:2103361 | NM_000548 | c.T244C | p.W82R | Missense | 0.48 |
| MCD_CCF_38 | M | 0-4 | 25-29 | MCD | FCD 2b | | *MTOR* | chr1:11217299 | NM_004958 | c.T4379C | p.L1460P | Missense | 0.03 |
| MCD_CCF_48 | F | 5-9 | 25-29 | MCD | FCD 2b | | *DEPDC5* | chr22:32161000 | NM_001242896 | c.G233C | p.R78P | Missense | 0.37 |
| MCD_CCF_48 | F | 5-9 | 25-29 | MCD | FCD 2b | | *TSC2* | chr16:2122364 | NM_000548 | c.G2220T | p.M740I | Missense | 0.48 |
| MCD_CCF_49 | F | 0-4 | 35-39 | MCD | FCD 2b | | *MTOR* | chr1:11188975 | NM_004958 | c.G5748T | p.W1916C | Missense | 0.46 |
| MCD_CCF_53 | M | 0-4 | 15-19 | MCD | FCD 2b | | *MTOR* | chr1:11169376 | NM_004958 | c.T7499A | p.I2500N | Missense | 0.04 |
| MCD_CCF_78 | M | 0-4 | 0-4 | MCD | FCD 2b | | *DEPDC5* | chr22:32200837 | NM_001242896 | c.C1153T | p.R385W | Missense | 0.43 |
| MCD_CCF_78 | M | 0-4 | 0-4 | MCD | FCD 2b | | *NPRL3* | chr16:188253 | ENST00000399953 | c.C14T | p.T5I | Missense | 0.55 |
| MCD_CCF_92 | M | 5-9 | 10-14 | MCD | FCD 2b | | *MTOR* | chr1:11187847 | NM_004958 | c.T6050C | p.I2017T | Missense | 0.08 |
| MCD_CCF_101 | F | 0-4 | 0-4 | MCD | FCD 2b | | *MTOR* | chr1:11184573 | NM_004958 | c.C6644A | p.S2215Y | Missense | 0.03 |
| MCD_CCF_102 | M | 5-9 | 15-19 | MCD | FCD 2b | | *MTOR* | chr1:11169377 | NM_004958 | c.A7498T | p.I2500F | Missense | 0.04 |
| MCD_CCF_102 | M | 5-9 | 15-19 | MCD | FCD 2b | | *TSC1* | chr9:135804196 | NM_000368 | c.C64T | p.R22W | Missense | 0.42 |
| MCD_EEBB_32 | M | 10-14 | 20-24 | MCD | FCD 2b | | *NF1* | chr17:29701091 | NM_001042492 | c.C8438T | p.T2813I | Missense | 0.21 |
| MCD_EEBB_67 | M | 20-24 | 30-34 | MCD | FCD 2b | | *NF1* | chr17:29677224 | NM_001042492 | c.G7345C | p.V2449L | Missense | 0.40 |
| MCD_EEBB_71 | F | 0-4 | 10-14 | MCD | FCD 2b | | *MTOR* | chr1:11188164 | NM_004958 | c.C5930A | p.T1977K | Missense | 0.05 |
| MCD_EEBB_80 | M | 5-9 | 15-19 | MCD | FCD 2b | | *MTOR* | chr1:11184573 | NM_004958 | c.C6644T | p.S2215F | Missense | 0.03 |
| MCD_EEBB_85 | F | 0-4 | 30-34 | MCD | FCD 2b | | *PTEN* | chr10:89623887 | NM_001304717 | c.180_181insAA | p.L61Nfs*2 | PTV | 0.49 |
| MCD_EEBB_92 | M | 0-4 | 0-4 | MCD | FCD 2b | | *MTOR* | chr1:11174399 | NM_004958 | c.7276delinsCCCT | p.P2425_L2426insP | Missense | 0.03 |
| MCD_CCF_93 | F | 0-4 | 0-4 | MCD | cMCD | HME / PMG / FCD 2a | *AKT3* | chr1:243859016 | NM_005465 | c.G49A | p.E17K | Missense | 0.08 |
| MCD_CCF_94 | F | 0-4 | 0-4 | MCD | cMCD | HME / PMG / FCD 2a / FCD 1a | *PTEN* | chr10:89720720 | NM_000314 | c.872dupA | p.N292Kfs*6 | PTV | 0.74 |

| | | | | | | | | | | | | | |
|---|---|---|---|---|---|---|---|---|---|---|---|---|---|
| MCD_CCF_110 | M | 0-4 | 0-4 | MCD | cMCD | HME / PMG / FCD 2a | *AKT3* | chr1:243859016 | NM_005465 | c.G49A | p.E17K | Missense | 0.13 |
| MCD_EEBB_14 | F | | 0-4 | MCD | cMCD | HME / FCD 2a | *MTOR* | chr1:11184573 | NM_004958 | c.C6644A | p.S2215Y | Missense | 0.10 |
| MCD_EEBB_21 | F | 0-4 | 0-4 | MCD | cMCD | HME / PMG / FCD 2a | *PIK3CA* | chr3:178936091 | NM_006218 | c.G1633A | p.E545K | Missense | 0.16 |
| MCD_EEBB_26 | F | 0-4 | 0-4 | MCD | cMCD | HME / PMG / FCD 2a | *PIK3CA* | chr3:178952085 | NM_006218 | c.A3140T | p.H1047L | Missense | 0.19 |
| MCD_EEBB_28 | M | 0-4 | 0-4 | MCD | cMCD | HME / FCD 2a | *MTOR* | chr1:11169377 | NM_004958 | c.A7498T | p.I2500F | Missense | 0.03 |
| MCD_EEBB_52 | F | 0-4 | 0-4 | MCD | cMCD | HME / PMG / FCD 2a | *PIK3CA* | chr3:178936082 | NM_006218 | c.G1624A | p.E542K | Missense | 0.27 |
| MCD_EEBB_57 | F | 0-4 | 0-4 | MCD | cMCD | HME / PMG / FCD 2a | *AKT3* | chr1:243859016 | NM_005465 | c.G49A | p.E17K | Missense | 0.08 |
| MCD_EEBB_57 | F | 0-4 | 0-4 | MCD | cMCD | HME / PMG / FCD 2a | *MTOR* | chr1:11313993 | NM_004958 | c.C743T | p.T248I | Missense | 0.47 |
| MCD_CCF_32 | M | 0-4 | 0-4 | MCD | cMCD | PMG / FCD 2a / FCD 1a | *NPRL3* | chr16:169168 | ENST00000399953 | c.G275A | p.R92Q | Missense | 0.30 |
| MCD_CCF_104 | F | 0-4 | 0-4 | MCD | cMCD | PMG / FCD 2b | *MTOR* | chr1:11301647 | NM_004958 | c.A1504C | p.K502Q | Missense | 0.09 |
| MCD_CCF_104 | F | 0-4 | 0-4 | MCD | cMCD | PMG / FCD 2b | *TSC2* | chr16:2135289 | NM_000548 | c.A4628G | p.H1543R | Missense | 0.41 |
| MCD_EEBB_10 | M | 10-14 | 25-29 | MCD | cMCD | PMG / NH / FCD 2a / GG | *NRAS* | chr1:115256529 | NM_002524 | c.A182G | p.Q61R | Missense | 0.13 |
| MCD_EEBB_65 | F | 0-4 | 20-24 | MCD | cMCD | PMG / NH / FCD 2a / DNET | *NRAS* | chr1:115256530 | NM_002524 | c.C181A | p.Q61K | Missense | 0.24 |
| MCD_EEBB_76 | M | 0-4 | 0-4 | MCD | cMCD | PMG / FCD 2b | *MTOR* | chr1:11174399 | NM_004958 | c.7276delinsCCCT | p.P2425_L2426insP | Missense | 0.04 |
| MCD_EEBB_89 | M | 5-9 | 20-24 | MCD | cMCD | PMG / FCD 1b | *NF1* | chr17:29483029 | NM_001042492 | c.C89A | p.T30K | Missense | 0.03 |
| MCD_EEBB_89 | M | 5-9 | 20-24 | MCD | cMCD | PMG / FCD 1b | *NF1* | chr17:29483016 | NM_001042492 | c.G76T | p.G26X | PTV | 0.46 |
| MCD_CCF_27 | F | 0-4 | 5-9 | MCD | TSC | FCD2b | *TSC2* | chr16:2130169 | NM_000548 | c.3402dupC | p.H1135Pfs*33 | PTV | 0.45 |
| MCD_CCF_29 | F | 0-4 | 0-4 | MCD | TSC | FCD 2b | *TSC2* | chr16:2121603 | NM_000548 | c.C1932A | p.C644X | PTV | 0.46 |
| MCD_CCF_55 | M | 0-4 | 5-9 | MCD | TSC | FCD2b | *TSC2* | chr16:2136739 | NM_000548 | c.T4856C | p.F1619S | Missense | 0.46 |
| MCD_CCF_56 | F | 0-4 | 5-9 | MCD | TSC | FCD2b | *TSC2* | chr16:2107157 | NM_000548 | c.826_827del | p.M276Vfs*61 | PTV | 0.43 |

Legend to supplementary table 2: HS = Hippocampal sclerosis, MCD = Malformation of cortical development, LEAT = Low-grade epilepsy-associated tumor, FCD = Focal cortical dysplasia, MOGHE = mild MCD with oligodendroglial hyperplasia in epilepsy, HME = Hemimegalencephaly, PMG = Polymicrogyria, cMCD = complex MCD, TSC = tuberous sclerosis complex, MA = meningioangiomatosis, DNET = Dysembryoplastic neuroepithelial tumor, PXA = Pleomorphic xanthoastrocytoma, MVNT = multinodular vacuolated neuronal tumor, SEGA = Subependymal giant cell astrocytoma, PTV = Protein-truncating variant

**Supplementary Table 5: Reported somatic CNV and CNN-LOH**

| Sample | Sex | Age at onset | Age at surgery | Lesion group | Pathology | Additional findings | Variant Position (hg19) | Region | Variant type | Included LFE genes | Size (Mbp) | Allelic fraction | Recurring | Samples w/ CNV |
|---|---|---|---|---|---|---|---|---|---|---|---|---|---|---|
| HS_EEBB_7 | M | 5-9 | 50-54 | HS | HS Type 1 | | 22:18520612-51304566 | 22q | CNN-LOH | *DEPDC5* | 32.8 | 0.03 | yes | 5 |
| HS_EEBB_81 | F | 0-4 | 25-29 | HS | HS Type 1 | FCD 3a | 19:50401320-59128983 | 19q13.42-13.43 | CNN-LOH | | 8.7 | 0.03 | yes | 2 |
| HS_EEBB_17 | M | 20-24 | 25-29 | HS | HS Type 2 | FCD 3a | 19:54462369-59128983 | 19q13.42-13.43 | CNN-LOH | | 4.7 | 0.02 | yes | 2 |
| HS_EEBB_55 | F | 15-19 | 45-49 | HS | HS (NOS) | | 9:86179563-141213431 | 9q21.33-34.2 | Loss | *TSC1* | 55.0 | 0.03 | yes | 2 |
| LEAT_EEBB_33 | M | 10-14 | 15-19 | LEAT | DNET | | 6:0-171115067 | 6 | Gain | *MYB* | 171.1 | 0.01 | yes | 8 |
| LEAT_EEBB_36 | M | 0-4 | 5-9 | LEAT | DNET | | 7:138562547-140335570 | 7q34 | Gain | | 1.8 | 0.20 | yes | 2 |
| LEAT_EEBB_41 | M | 0-4 | 0-4 | LEAT | DNET | | 8:0-38684853 | 8p | CNN-LOH | *FGFR1* | 38.7 | 0.11 | yes | 2 |
| LEAT_EEBB_49 | F | 30-34 | 30-34 | LEAT | DNET | | 19:44544237-59128983 | 19q13.31-13.4 | Loss | | 14.6 | 0.05 | no | 1 |
| LEAT_EEBB_49 | F | 30-34 | 30-34 | LEAT | DNET | | 20:0-22746255 | 20p | Loss | | 22.7 | 0.03 | no | 1 |
| LEAT_EEBB_55 | M | 20-24 | 25-29 | LEAT | DNET | | 2:25568821-63939417 | 2p23.3-15 | Gain | | 38.4 | 0.21 | no | 1 |
| LEAT_EEBB_55 | M | 20-24 | 25-29 | LEAT | DNET | | 2:0-25424397 | 2p25.3-23.3 | Loss | | 25.4 | 0.26 | no | 1 |
| LEAT_EEBB_55 | M | 20-24 | 25-29 | LEAT | DNET | | 8:49729020-94305178 | 8q11.21-22.1 | Loss | *MYBL1* | 44.6 | 0.11 | no | 1 |
| LEAT_EEBB_55 | M | 20-24 | 25-29 | LEAT | DNET | | 13:28150993-115169878 | 13 | Loss | | 87.0 | 0.26 | yes | 2 |
| LEAT_EEBB_55 | M | 20-24 | 25-29 | LEAT | DNET | | 15:23833737-60664030 | 15q11.2-22.2 | Loss | | 36.8 | 0.05 | no | 1 |
| LEAT_EEBB_75 | M | 5-9 | 10-14 | LEAT | DNET | | 2:0-243199373 | 2 | Gain | | 243.2 | 0.01 | yes | 3 |
| LEAT_EEBB_75 | M | 5-9 | 10-14 | LEAT | DNET | | 7:0-147731239 | 7 | Gain | *BRAF* | 147.7 | 0.02 | yes | 18 |
| LEAT_EEBB_75 | M | 5-9 | 10-14 | LEAT | DNET | | 7:138550994-140481402 | 7q34 | Gain | *BRAF* | 1.9 | 0.30 | yes | 2 |
| LEAT_EEBB_75 | M | 5-9 | 10-14 | LEAT | DNET | | 12:49424065-51343851 | 12q13.12 | Gain | | 1.9 | 0.32 | no | 1 |
| LEAT_EEBB_81 | M | 45-49 | 55-59 | LEAT | DNET | | 8:0-42446238 | 8p | CNN-LOH | *FGFR1* | 42.4 | 0.04 | yes | 2 |
| LEAT_EEBB_104 | M | 5-9 | 15-19 | LEAT | DNET | | 4:0-191154276 | 4 | Gain | | 191.2 | 0.12 | yes | 5 |
| LEAT_EEBB_104 | M | 5-9 | 15-19 | LEAT | DNET | | 6:0-171115067 | 6 | Gain | *MYB* | 171.1 | 0.11 | yes | 8 |
| LEAT_EEBB_104 | M | 5-9 | 15-19 | LEAT | DNET | | 7:0-159138663 | 7 | Gain | *RHEB, BRAF* | 159.1 | 0.12 | yes | 18 |
| LEAT_EEBB_104 | M | 5-9 | 15-19 | LEAT | DNET | | 9:0-141213431 | 9 | Gain | *TSC1* | 141.2 | 0.12 | yes | 2 |
| LEAT_EEBB_104 | M | 5-9 | 15-19 | LEAT | DNET | | 11:0-135006516 | 11 | Gain | | 135.0 | 0.13 | yes | 5 |

| | | | | | | | | | | | | | | |
|---|---|---|---|---|---|---|---|---|---|---|---|---|---|---|
| LEAT_EEBB_104 | M | 5-9 | 15-19 | LEAT | DNET | | 14:1.9e+07-107349540 | 14q | Gain | | 88.3 | 0.12 | yes | 2 |
| LEAT_EEBB_104 | M | 5-9 | 15-19 | LEAT | DNET | | 20:0-63025520 | 20 | Gain | | 63.0 | 0.12 | yes | 10 |
| LEAT_EEBB_105 | F | 35-39 | 35-39 | LEAT | DNET | | 6:0-171115067 | 6 | Gain | *MYB* | 171.1 | 0.02 | yes | 8 |
| LEAT_EEBB_105 | F | 35-39 | 35-39 | LEAT | DNET | | 7:0-159138663 | 7 | Gain | *RHEB, BRAF* | 159.1 | 0.08 | yes | 18 |
| LEAT_EEBB_1 | F | 10-14 | 15-19 | LEAT | Ganglioglioma | | 5:0-180915260 | 5 | Gain | | 180.9 | 0.20 | yes | 11 |
| LEAT_EEBB_1 | F | 10-14 | 15-19 | LEAT | Ganglioglioma | | 7:0-159138663 | 7 | Gain | *RHEB, BRAF* | 159.1 | 0.21 | yes | 18 |
| LEAT_EEBB_1 | F | 10-14 | 15-19 | LEAT | Ganglioglioma | | 11:0-135006516 | 11 | Gain | | 135.0 | 0.20 | yes | 5 |
| LEAT_EEBB_1 | F | 10-14 | 15-19 | LEAT | Ganglioglioma | | 12:0-133851895 | 12 | Gain | *KRAS, PTPN11* | 133.9 | 0.20 | yes | 8 |
| LEAT_EEBB_1 | F | 10-14 | 15-19 | LEAT | Ganglioglioma | | 19:0-59128983 | 19 | Gain | | 59.1 | 0.20 | yes | 5 |
| LEAT_EEBB_1 | F | 10-14 | 15-19 | LEAT | Ganglioglioma | | 20:0-63025520 | 20 | Gain | | 63.0 | 0.19 | yes | 10 |
| LEAT_EEBB_4 | M | 0-4 | 15-19 | LEAT | Ganglioglioma | | 10:118766183-123216161 | 10q25.3-26.13 | Loss | | 4.4 | 0.22 | no | 1 |
| LEAT_EEBB_11 | F | 5-9 | 10-14 | LEAT | Ganglioglioma | | 1:20142866-249250621 | 1 | Gain | *AKT3* | 229.1 | 0.03 | no | 1 |
| LEAT_EEBB_11 | F | 5-9 | 10-14 | LEAT | Ganglioglioma | | 5:0-180915260 | 5 | Gain | | 180.9 | 0.03 | yes | 11 |
| LEAT_EEBB_11 | F | 5-9 | 10-14 | LEAT | Ganglioglioma | | 20:921971-63025520 | 20 | Gain | | 62.1 | 0.07 | yes | 10 |
| LEAT_EEBB_15 | M | 15-19 | 15-19 | LEAT | Ganglioglioma | HS | 9:0-141213431 | 9 | Loss | *TSC1* | 141.2 | 0.06 | yes | 3 |
| LEAT_EEBB_16 | M | 5-9 | 20-24 | LEAT | Ganglioglioma | | 3:0-56942973 | 3p26.3-14.3 | Loss | *NPRL2, RAF1* | 56.9 | 0.04 | no | 1 |
| LEAT_EEBB_16 | M | 5-9 | 20-24 | LEAT | Ganglioglioma | | 22:17725150-51304566 | 22q | Loss | *DEPDC5* | 33.6 | 0.04 | yes | 4 |
| LEAT_EEBB_32 | F | 20-24 | 20-24 | LEAT | Ganglioglioma | | 5:0-180915260 | 5 | Gain | | 180.9 | 0.29 | yes | 11 |
| LEAT_EEBB_32 | F | 20-24 | 20-24 | LEAT | Ganglioglioma | | 7:0-159138663 | 7 | Gain | *RHEB, BRAF* | 159.1 | 0.29 | yes | 18 |
| LEAT_EEBB_32 | F | 20-24 | 20-24 | LEAT | Ganglioglioma | | 12:0-133851895 | 12 | Gain | *KRAS, PTPN11* | 133.9 | 0.29 | yes | 8 |
| LEAT_EEBB_40 | F | 10-14 | 25-29 | LEAT | Ganglioglioma | HS Type 1 | 19:0-59128983 | 19 | Gain | | 59.1 | 0.18 | yes | 5 |
| LEAT_EEBB_40 | F | 10-14 | 25-29 | LEAT | Ganglioglioma | HS Type 1 | 20:0-63025520 | 20 | Gain | | 63.0 | 0.19 | yes | 10 |
| LEAT_EEBB_42 | F | 20-24 | 30-34 | LEAT | Ganglioglioma | | 21:15515843-48129895 | 21q | CNN-LOH | | 32.6 | 0.05 | yes | 2 |
| LEAT_EEBB_42 | F | 20-24 | 30-34 | LEAT | Ganglioglioma | | 22:17221495-51304566 | 22q | CNN-LOH | *DEPDC5* | 34.1 | 0.06 | yes | 5 |
| LEAT_EEBB_42 | F | 20-24 | 30-34 | LEAT | Ganglioglioma | | 2:0-243199373 | 2 | Gain | | 243.2 | 0.07 | yes | 3 |
| LEAT_EEBB_42 | F | 20-24 | 30-34 | LEAT | Ganglioglioma | | 5:0-180915260 | 5 | Gain | | 180.9 | 0.05 | yes | 11 |

| | | | | | | | | | | | | | |
|---|---|---|---|---|---|---|---|---|---|---|---|---|---|
| LEAT_EEBB_42 | F | 20-24 | 30-34 | LEAT | Ganglioglioma | | 7:0-159138663 | 7 | Gain | *RHEB, BRAF* | 159.1 | 0.04 | yes | 18 |
| LEAT_EEBB_42 | F | 20-24 | 30-34 | LEAT | Ganglioglioma | | 12:0-133851895 | 12 | Gain | *KRAS, PTPN11* | 133.9 | 0.07 | yes | 8 |
| LEAT_EEBB_42 | F | 20-24 | 30-34 | LEAT | Ganglioglioma | | 14:24860592-107349540 | 14q | Gain | | 82.5 | 0.06 | yes | 2 |
| LEAT_EEBB_42 | F | 20-24 | 30-34 | LEAT | Ganglioglioma | | 15:23226254-102531392 | 15q | Gain | | 79.3 | 0.04 | yes | 3 |
| LEAT_EEBB_48 | M | 25-29 | 40-44 | LEAT | Ganglioglioma | HS Type 1 | 4:58377516-191154276 | 4q | Gain | | 132.8 | 0.02 | no | 1 |
| LEAT_EEBB_63 | M | 10-14 | 15-19 | LEAT | Ganglioglioma | | 12:0-133851895 | 12 | Gain | *KRAS, PTPN11* | 133.9 | 0.05 | yes | 8 |
| LEAT_EEBB_63 | M | 10-14 | 15-19 | LEAT | Ganglioglioma | | 18:0-78077248 | 18 | Gain | | 78.1 | 0.05 | no | 1 |
| LEAT_EEBB_63 | M | 10-14 | 15-19 | LEAT | Ganglioglioma | | 19:6547141-59128983 | 19 | Gain | | 52.6 | 0.05 | yes | 5 |
| LEAT_EEBB_63 | M | 10-14 | 15-19 | LEAT | Ganglioglioma | | 20:0-63025520 | 20 | Gain | | 63.0 | 0.05 | yes | 10 |
| LEAT_EEBB_63 | M | 10-14 | 15-19 | LEAT | Ganglioglioma | | 13:21050575-115169878 | 13 | Loss | | 94.1 | 0.05 | yes | 2 |
| LEAT_EEBB_70 | F | 15-19 | 20-24 | LEAT | Ganglioglioma | | 5:0-180915260 | 5 | Gain | | 180.9 | 0.05 | yes | 11 |
| LEAT_EEBB_70 | F | 15-19 | 20-24 | LEAT | Ganglioglioma | | 7:0-159138663 | 7 | Gain | *RHEB, BRAF* | 159.1 | 0.05 | yes | 18 |
| LEAT_EEBB_70 | F | 15-19 | 20-24 | LEAT | Ganglioglioma | | 11:0-135006516 | 11 | Gain | | 135.0 | 0.11 | yes | 5 |
| LEAT_EEBB_70 | F | 15-19 | 20-24 | LEAT | Ganglioglioma | | 13:19307875-115169878 | 13 | Gain | | 95.9 | 0.11 | yes | 2 |
| LEAT_EEBB_74 | M | 5-9 | 10-14 | LEAT | Ganglioglioma | | 4:0-191154276 | 4 | Gain | | 191.2 | 0.09 | yes | 5 |
| LEAT_EEBB_74 | M | 5-9 | 10-14 | LEAT | Ganglioglioma | | 6:0-171115067 | 6 | Gain | *MYB* | 171.1 | 0.06 | yes | 8 |
| LEAT_EEBB_74 | M | 5-9 | 10-14 | LEAT | Ganglioglioma | | 7:0-159138663 | 7 | Gain | *RHEB, BRAF* | 159.1 | 0.06 | yes | 18 |
| LEAT_EEBB_74 | M | 5-9 | 10-14 | LEAT | Ganglioglioma | | 9:0-141213431 | 9 | Gain | *TSC1* | 141.2 | 0.08 | yes | 2 |
| LEAT_EEBB_74 | M | 5-9 | 10-14 | LEAT | Ganglioglioma | | 12:0-133851895 | 12 | Gain | *KRAS, PTPN11* | 133.9 | 0.06 | yes | 8 |
| LEAT_EEBB_74 | M | 5-9 | 10-14 | LEAT | Ganglioglioma | | 16:0-90354753 | 16 | Gain | *NPRL3, TSC2* | 90.4 | 0.08 | yes | 3 |
| LEAT_EEBB_74 | M | 5-9 | 10-14 | LEAT | Ganglioglioma | | 20:0-63025520 | 20 | Gain | | 63.0 | 0.06 | yes | 10 |
| LEAT_EEBB_80 | M | 5-9 | 5-9 | LEAT | Ganglioglioma | | 5:0-180915260 | 5 | Gain | | 180.9 | 0.03 | yes | 11 |
| LEAT_EEBB_80 | M | 5-9 | 5-9 | LEAT | Ganglioglioma | | 6:0-171115067 | 6 | Gain | *MYB* | 171.1 | 0.03 | yes | 8 |
| LEAT_EEBB_80 | M | 5-9 | 5-9 | LEAT | Ganglioglioma | | 7:0-159138663 | 7 | Gain | *RHEB, BRAF* | 159.1 | 0.03 | yes | 18 |
| LEAT_EEBB_80 | M | 5-9 | 5-9 | LEAT | Ganglioglioma | | 19:0-23207354 | 19p | Gain | | 23.2 | 0.12 | no | 1 |
| LEAT_EEBB_80 | M | 5-9 | 5-9 | LEAT | Ganglioglioma | | 1:0-120110175 | 1p | Loss | *MTOR* | 120.1 | 0.08 | no | 1 |

| | | | | | | | | | | | | | | |
|---|---|---|---|---|---|---|---|---|---|---|---|---|---|---|
| LEAT_EEBB_80 | M | 5-9 | 5-9 | LEAT | Ganglioglioma | | 22:17897544-51304566 | 22q | Loss | *DEPDC5* | 33.4 | 0.06 | yes | 4 |
| LEAT_EEBB_85 | M | | 30-34 | LEAT | Ganglioglioma | | 4:0-191154276 | 4 | Gain | | 191.2 | 0.04 | yes | 5 |
| LEAT_EEBB_85 | M | | 30-34 | LEAT | Ganglioglioma | | 6:0-171115067 | 6 | Gain | *MYB* | 171.1 | 0.04 | yes | 8 |
| LEAT_EEBB_87 | F | 10-14 | 50-54 | LEAT | Ganglioglioma | | 5:0-180915260 | 5 | Gain | | 180.9 | 0.04 | yes | 11 |
| LEAT_EEBB_87 | F | 10-14 | 50-54 | LEAT | Ganglioglioma | | 7:0-159138663 | 7 | Gain | *RHEB, BRAF* | 159.1 | 0.04 | yes | 18 |
| LEAT_EEBB_87 | F | 10-14 | 50-54 | LEAT | Ganglioglioma | | 12:0-133851895 | 12 | Gain | *KRAS, PTPN11* | 133.9 | 0.04 | yes | 8 |
| LEAT_EEBB_87 | F | 10-14 | 50-54 | LEAT | Ganglioglioma | | 20:0-63025520 | 20 | Gain | | 63.0 | 0.02 | yes | 10 |
| LEAT_EEBB_98 | M | 15-19 | 15-19 | LEAT | Ganglioglioma | | 18:18570235-78077248 | 18q | CNN-LOH | | 59.5 | 0.01 | no | 1 |
| LEAT_EEBB_98 | M | 15-19 | 15-19 | LEAT | Ganglioglioma | | 21:20190260-48129895 | 21q | CNN-LOH | | 27.9 | 0.03 | yes | 2 |
| LEAT_EEBB_98 | M | 15-19 | 15-19 | LEAT | Ganglioglioma | | 4:0-191154276 | 4 | Gain | | 191.2 | 0.02 | yes | 5 |
| LEAT_EEBB_98 | M | 15-19 | 15-19 | LEAT | Ganglioglioma | | 7:0-134639161 | 7 | Gain | | 134.6 | 0.03 | yes | 18 |
| LEAT_EEBB_98 | M | 15-19 | 15-19 | LEAT | Ganglioglioma | | 11:0-135006516 | 11 | Gain | | 135.0 | 0.04 | yes | 5 |
| LEAT_EEBB_101 | F | 5-9 | 10-14 | LEAT | Ganglioglioma | | 5:0-180915260 | 5 | Gain | | 180.9 | 0.29 | yes | 11 |
| LEAT_EEBB_101 | F | 5-9 | 10-14 | LEAT | Ganglioglioma | | 6:0-171115067 | 6 | Gain | *MYB* | 171.1 | 0.33 | yes | 8 |
| LEAT_EEBB_101 | F | 5-9 | 10-14 | LEAT | Ganglioglioma | | 7:0-159138663 | 7 | Gain | *RHEB, BRAF* | 159.1 | 0.33 | yes | 18 |
| LEAT_EEBB_101 | F | 5-9 | 10-14 | LEAT | Ganglioglioma | | 10:0-135269349 | 10 | Gain | *PTEN* | 135.3 | 0.32 | yes | 3 |
| LEAT_EEBB_101 | F | 5-9 | 10-14 | LEAT | Ganglioglioma | | 15:20052938-102531392 | 15q | Gain | | 82.5 | 0.32 | yes | 3 |
| LEAT_EEBB_101 | F | 5-9 | 10-14 | LEAT | Ganglioglioma | | 20:0-63025520 | 20 | Gain | | 63.0 | 0.32 | yes | 10 |
| LEAT_EEBB_101 | F | 5-9 | 10-14 | LEAT | Ganglioglioma | | 22:16000000-51304566 | 22q | Gain | *DEPDC5* | 35.3 | 0.31 | no | 1 |
| LEAT_EEBB_103 | F | 15-19 | 20-24 | LEAT | Ganglioglioma | HS Type 1 | 2:0-243199373 | 2 | Gain | | 243.2 | 0.02 | yes | 3 |
| LEAT_EEBB_103 | F | 15-19 | 20-24 | LEAT | Ganglioglioma | HS Type 1 | 3:0-188200897 | 3 | Gain | *NPRL2, RAF1, PIK3CA* | 188.2 | 0.02 | no | 1 |
| LEAT_EEBB_103 | F | 15-19 | 20-24 | LEAT | Ganglioglioma | HS Type 1 | 7:0-159138663 | 7 | Gain | *RHEB, BRAF* | 159.1 | 0.27 | yes | 18 |
| LEAT_EEBB_103 | F | 15-19 | 20-24 | LEAT | Ganglioglioma | HS Type 1 | 16:0-90354753 | 16 | Gain | *NPRL3, TSC2* | 90.4 | 0.02 | yes | 3 |
| LEAT_EEBB_103 | F | 15-19 | 20-24 | LEAT | Ganglioglioma | HS Type 1 | 9:0-141213431 | 9 | Loss | *TSC1* | 141.2 | 0.36 | yes | 3 |
| LEAT_EEBB_103 | F | 15-19 | 20-24 | LEAT | Ganglioglioma | HS Type 1 | 17:0-75075664 | 17 | Loss | *NF1* | 75.1 | 0.05 | no | 1 |
| LEAT_EEBB_103 | F | 15-19 | 20-24 | LEAT | Ganglioglioma | HS Type 1 | 22:17223938-51304566 | 22q | Loss | *DEPDC5* | 34.1 | 0.08 | yes | 4 |

| | | | | | | | | | | | | | | |
|---|---|---|---|---|---|---|---|---|---|---|---|---|---|---|
| LEAT_EEBB_110 | M | 5-9 | 15-19 | LEAT | Angiocentric glioma | | 6:135543837-163969815 | 6q23.3-24.1 | Loss | | 28.4 | 0.20 | yes | 2 |
| LEAT_EEBB_46 | M | 10-14 | 20-24 | LEAT | Angiocentric glioma | | 6:135521495-141454143 | 6q23.3-24.1 | Loss | *MYB* | 5.9 | 0.69 | yes | 2 |
| LEAT_EEBB_19 | F | 15-19 | 15-19 | LEAT | Desmoplastic Ganglioglioma | | 5:71661131-180915260 | 5q13.2-35.3 | Loss | | 109.3 | 0.23 | no | 1 |
| LEAT_EEBB_19 | F | 15-19 | 15-19 | LEAT | Desmoplastic Ganglioglioma | | 9:73909871-86175639 | 9q21.12-21.32 | Loss | | 12.3 | 0.22 | no | 1 |
| LEAT_EEBB_19 | F | 15-19 | 15-19 | LEAT | Desmoplastic Ganglioglioma | | 9:88234954-137530346 | 9q21.33-34.2 | Loss | *TSC1* | 49.3 | 0.23 | yes | 2 |
| LEAT_EEBB_19 | F | 15-19 | 15-19 | LEAT | Desmoplastic Ganglioglioma | | 22:23754578-51304566 | 22q | Loss | *DEPDC5* | 27.5 | 0.22 | yes | 4 |
| LEAT_EEBB_112 | M | 5-9 | 25-29 | LEAT | IDA | | 9:0-141213431 | 9 | Loss | *TSC1* | 141.2 | 0.22 | yes | 3 |
| LEAT_EEBB_112 | M | 5-9 | 25-29 | LEAT | IDA | | 18:0-78077248 | 18 | Loss | | 78.1 | 0.24 | no | 1 |
| LEAT_EEBB_50 | M | 25-29 | 25-29 | LEAT | IDA | HS Type 1 | 5:0-180915260 | 5 | Gain | | 180.9 | 0.35 | yes | 11 |
| LEAT_EEBB_50 | M | 25-29 | 25-29 | LEAT | IDA | HS Type 1 | 7:0-159138663 | 7 | Gain | *RHEB, BRAF* | 159.1 | 0.35 | yes | 18 |
| LEAT_EEBB_50 | M | 25-29 | 25-29 | LEAT | IDA | HS Type 1 | 11:0-135006516 | 11 | Gain | | 135.0 | 0.34 | yes | 5 |
| LEAT_EEBB_50 | M | 25-29 | 25-29 | LEAT | IDA | HS Type 1 | 12:0-133851895 | 12 | Gain | *KRAS, PTPN11* | 133.9 | 0.06 | yes | 8 |
| LEAT_EEBB_50 | M | 25-29 | 25-29 | LEAT | IDA | HS Type 1 | 13:19377679-115169878 | 13 | Gain | | 95.8 | 0.06 | yes | 2 |
| LEAT_EEBB_50 | M | 25-29 | 25-29 | LEAT | IDA | HS Type 1 | 17:0-81195210 | 17 | Gain | *NF1* | 81.2 | 0.24 | no | 1 |
| LEAT_EEBB_50 | M | 25-29 | 25-29 | LEAT | IDA | HS Type 1 | 19:0-59128983 | 19 | Gain | | 59.1 | 0.33 | yes | 5 |
| LEAT_EEBB_50 | M | 25-29 | 25-29 | LEAT | IDA | HS Type 1 | 20:0-63025520 | 20 | Gain | | 63.0 | 0.03 | yes | 10 |
| LEAT_EEBB_113 | M | 0-4 | 44-49 | LEAT | MVNT | | 5:0-180915260 | 5 | Gain | | 180.9 | 0.02 | yes | 11 |
| LEAT_EEBB_113 | M | 0-4 | 44-49 | LEAT | MVNT | | 7:0-159138663 | 7 | Gain | *RHEB, BRAF* | 159.1 | 0.06 | yes | 18 |
| LEAT_EEBB_102 | F | 5-9 | 10-14 | LEAT | Pilocytic Astrocytoma | | 17:52121998-81195210 | 17q22-25.3 | Gain | | 29.1 | 0.23 | no | 1 |
| LEAT_EEBB_95 | F | 5-9 | 34-39 | LEAT | Pilocytic Astrocytoma | | 17:26851501-81195210 | 17q | CNN-LOH | *NF1* | 54.3 | 0.12 | yes | 2 |
| LEAT_EEBB_95 | F | 5-9 | 34-39 | LEAT | Pilocytic Astrocytoma | | 4:0-191154276 | 4 | Gain | | 191.2 | 0.15 | yes | 5 |
| LEAT_EEBB_95 | F | 5-9 | 34-39 | LEAT | Pilocytic Astrocytoma | | 6:0-171115067 | 6 | Gain | *MYB* | 171.1 | 0.06 | yes | 8 |
| LEAT_EEBB_95 | F | 5-9 | 34-39 | LEAT | Pilocytic Astrocytoma | | 7:0-159138663 | 7 | Gain | *RHEB, BRAF* | 159.1 | 0.15 | yes | 18 |
| LEAT_EEBB_95 | F | 5-9 | 34-39 | LEAT | Pilocytic Astrocytoma | | 10:0-135534747 | 10 | Gain | *PTEN* | 135.5 | 0.14 | yes | 3 |
| LEAT_EEBB_95 | F | 5-9 | 34-39 | LEAT | Pilocytic Astrocytoma | | 19:0-59128983 | 19 | Gain | | 59.1 | 0.07 | yes | 5 |
| LEAT_EEBB_56 | M | 10-14 | 15-19 | LEAT | PXA | | 5:0-180915260 | 5 | Gain | | 180.9 | 0.10 | yes | 11 |

| | | | | | | | | | | | | | | |
|---|---|---|---|---|---|---|---|---|---|---|---|---|---|---|
| LEAT_EEBB_56 | M | 10-14 | 15-19 | LEAT | PXA | | 7:0-65796278 | 7p22.3-q11.21 | Gain | | 65.8 | 0.11 | no | 1 |
| LEAT_EEBB_56 | M | 10-14 | 15-19 | LEAT | PXA | | 10:0-135534747 | 10 | Gain | *PTEN* | 135.5 | 0.12 | yes | 3 |
| LEAT_EEBB_56 | M | 10-14 | 15-19 | LEAT | PXA | | 12:0-133851895 | 12 | Gain | *KRAS, PTPN11* | 133.9 | 0.12 | yes | 8 |
| LEAT_EEBB_56 | M | 10-14 | 15-19 | LEAT | PXA | | 15:24148457-102531392 | 15q | Gain | | 78.4 | 0.12 | yes | 3 |
| LEAT_EEBB_56 | M | 10-14 | 15-19 | LEAT | PXA | | 16:0-90354753 | 16 | Gain | *NPRL3, TSC2* | 90.4 | 0.12 | yes | 3 |
| LEAT_EEBB_56 | M | 10-14 | 15-19 | LEAT | PXA | | 20:0-63025520 | 20 | Gain | | 63.0 | 0.12 | yes | 10 |
| LEAT_EEBB_56 | M | 10-14 | 15-19 | LEAT | PXA | | 21:15463956-48129895 | 21q | Gain | | 32.7 | 0.14 | no | 1 |
| LEAT_EEBB_56 | M | 10-14 | 15-19 | LEAT | PXA | | 7:65796279-159138663 | 7q | Loss | *RHEB, BRAF* | 93.3 | 0.24 | no | 1 |
| LEAT_CCF_37 | M | 0-4 | 5-9 | LEAT | TSC | SEGA | 16:0-9806375 | 16p13.3 | CNN-LOH | *NPRL3, TSC2* | 9.8 | 0.03 | yes | 4 |
| LEAT_CCF_37 | M | 0-4 | 5-9 | LEAT | TSC | SEGA | 7:0-159138663 | 7 | Gain | *RHEB, BRAF* | 159.1 | 0.14 | yes | 18 |
| LEAT_EEBB_17 | F | 0-4 | 5-9 | LEAT | TSC | SEGA | 16:0-16081737 | 16p13.3 | CNN-LOH | *NPRL3, TSC2* | 16.1 | 0.09 | yes | 4 |
| MCD_CCF_62 | F | 5-9 | 5-9 | MCD | FCD 2a | Hyaline astrocytic inclusions | 1:146523045-249250621 | 1q | Gain | *AKT3* | 102.7 | 0.06 | yes | 2 |
| MCD_CCF_62 | F | 5-9 | 5-9 | MCD | FCD 2a | Hyaline astrocytic inclusions | 16:47304567-90354753 | 16q12.1-24.3 | Loss | | 43.1 | 0.06 | no | 1 |
| MCD_EEBB_68 | M | 5-9 | 40-44 | MCD | FCD 2a | | 22:18009909-49085205 | 22q | CNN-LOH | *DEPDC5* | 31.1 | 0.04 | yes | 5 |
| MCD_EEBB_75 | M | 0-4 | 10-14 | MCD | FCD 2a | | 22:19183787-51304566 | 22q | CNN-LOH | *DEPDC5* | 32.1 | 0.04 | yes | 5 |
| MCD_EEBB_94 | F | 0-4 | 0-4 | MCD | FCD 2a | | 22:1.6e+07-51304566 | 22q | CNN-LOH | *DEPDC5* | 35.3 | 0.04 | yes | 5 |
| MCD_EEBB_110 | M | 0-4 | 5-9 | MCD | FCD 2a | Hyaline astrocytic inclusions | 1:145723739-249250621 | 1q | Gain | *AKT3* | 103.5 | 0.08 | yes | 2 |
| MCD_CCF_48 | F | 5-9 | 25-29 | MCD | FCD 2b | | 9:71034203-141213431 | 9q21.11-34.3 | CNN-LOH | *TSC1* | 70.2 | 0.09 | no | 1 |
| MCD_CCF_48 | F | 5-9 | 25-29 | MCD | FCD 2b | | 16:0-3367061 | 16p13.3 | CNN-LOH | *NPRL3, TSC2* | 3.4 | 0.05 | yes | 4 |
| MCD_CCF_90 | F | 0-4 | 0-4 | MCD | FCD 2b | | 7:0-159138663 | 7 | Gain | *RHEB, BRAF* | 159.1 | 0.05 | yes | 18 |
| MCD_EEBB_104 | F | | 15-19 | MCD | FCD 2b | | 4:93918343-191154276 | 4q22.2-35.2 | CNN-LOH | | 97.2 | 0.02 | no | 1 |
| MCD_CCF_71 | F | 0-4 | 0-4 | MCD | cMCD | PMG / NH | 1:203644628-234525940 | 1q32.1-42.2 | Gain | | 30.9 | 0.06 | no | 1 |
| MCD_CCF_94 | F | 0-4 | 0-4 | MCD | cMCD | HME / PMG / FCD 2a / FCD 1a | 10:75324903-135534747 | 10q22.2-26.3 | CNN-LOH | *PTEN* | 60.2 | 0.15 | no | 1 |
| MCD_CCF_104 | F | 0-4 | 0-4 | MCD | cMCD | PMG / FCD 2b | 16:0-12353187 | 16p13.3 | CNN-LOH | *NPRL3, TSC2* | 12.4 | 0.04 | yes | 4 |
| MCD_EEBB_89 | M | 5-9 | 20-24 | MCD | cMCD | PMG / FCD 1b | 17:28305017-81195210 | 17q | CNN-LOH | *NF1* | 52.9 | 0.06 | yes | 2 |

Legend to supplementary table 3: LFE = lesional focal epilepsy, CNV = Copy number variant, LEAT = Low-grade epilepsy-associated tumor, MCD = Malformation of cortical development, HS = Hippocampal sclerosis, DNET = Dysembryoplastic neuroepithelial tumor, IDA = Isomorphic diffuse astrocytoma, PXA = Pleomorphic xanthoastrocytoma, MVNT = multinodular vacuolated neuronal tumor, SEGA = Subependymal giant cell astrocytoma, TSC = Tuberous sclerosis complex, FCD = Focal cortical dysplasia, HME = Hemimegalencephaly, cMCD = complex MCD, PMG = Polymicrogyria, CNN-LOH = Copy number neutral loss of heterozygosity
